# Exploring the Privacy-Utility Tradeoff in Differentially Private Federated Learning for Mobile Health: A Novel Approach using Simulated Privacy Attacks

**DOI:** 10.1101/2022.10.17.22281116

**Authors:** Alexander Shen, Luke Francisco, Srijan Sen, Ambuj Tewari

## Abstract

**Background:** While evidence supporting the feasibility of large scale mHealth systems continues to grow, privacy protection continues to be an important implementation challenge. The potential scale of publicly available mHealth applications and the sensitive nature of the data involved will inevitably attract unwanted attention from adversarial actors seeking to compromise user privacy. Although privacy-preserving technologies such as Federated Learning and Differential Privacy offers strong theoretical guarantees, it is not clear how such technologies actually perform under real-world conditions.

**Objective:** Using data from the University of Michigan Intern Health Study (IHS), we assess the privacy protection capabilities of Federated Learning and Differential Privacy against the associated tradeoffs in model accuracy and training time using simulation methods. Specifically, our objectives are to (1) construct a “target” mHealth system using the demographic and sensor data available in the IHS (2) mount a simulated privacy attack that attempts to compromise IHS participant privacy (3) measure the effectiveness of such an attack under various levels of privacy protection on the target mHealth system, and (4) measure the costs to algorithmic performance associated with the chosen levels of privacy protection.

**Methods:** For (1), we perform simple data processing/imputation and construct a neural network classifier that attempts to predict participant daily mood EMA score from sensor data. For (2) we make certain assumptions of the attacker’s capabilities and construct an attack intended to uncover statistical properties of private participant data based on techniques proposed in the literature. For (3) and (4), we report a collection of conventional metrics to evaluate the success of the privacy attack and performance of the original mHealth system under Federated Learning and various levels of Differential Privacy.

**Results:** We find that Federated Learning alone does not provide adequate protection against the privacy attack proposed above, where the attacker’s success rate in identifying private data attributes is over 90% in the worst case. However, under the highest level of Differential Privacy tested in this paper, the attacker’s success rate falls to around 59.6% with only a 10 percentage point decrease in model R^2^ and a 42% increase in model training time. Finally, we show that those participants in the IHS most likely to require strong privacy protection are also most at risk from this particular privacy attack and subsequently stand to benefit the most from these privacy-preserving technologies.

**Conclusions:** Our results demonstrate both the necessity of proactive privacy protection research and the feasibility of current Federated Learning and Differential Privacy methods implemented in a real mHealth scenario. Our simulation methods for privacy protection measurement provide a novel framework for characterizing the privacy-utility tradeoff and serve as a potential foundation for future research.

## Introduction

The rise of Mobile Health (mHealth) as an exciting and compelling healthcare paradigm has been unmistakable. As wearable devices continue to gain popularity and smartphone penetration continues to rise in developing countries[1][2], opportunities for mHealth to make positive impacts in healthcare delivery and administration have proliferated. On the research side, 2018-2020 has seen more mHealth related publications than all previous years combined[3]. Meanwhile, the current global mHealth market is expected to experience annual growth of 11.0% for the better part of this decade[2].

Still, widespread adoption of mHealth technologies may not occur until the challenge of user data privacy is overcome[4]. Recent polling has shown that the vast majority of Americans do not believe that the benefits of releasing personal data to private companies are worth the risks[5]. Research has also shown that privacy concerns directly affect the willingness of users to participate in mHealth systems, especially for younger users[6] and users who suffer from diseases that carry social stigma[7]. Along with the increased frequency of cyber attacks on centralized data centers/IT infrastructure and the growing number of smart devices that utilize these facilities[8], the costs of insufficient security and privacy protection for future large-scale mHealth applications are abundantly clear.

In this paper, we examine two popular privacy protection strategies (Federated Learning and Differential Privacy) and explore some of their shortcomings. Specifically, we demonstrate the tradeoff between algorithmic utility and privacy protection when implementing these strategies in practical mHealth settings. We hope to offer insight into the potential of these strategies to protect user data by constructing a simulated attack on a centralized server.

Federated Learning

Federated Learning (FL) is a machine learning method used when data is distributed across independent devices (often referred to as “clients”). In the traditional “Centralized Learning” regime, a data collector interested in constructing a statistical model for answering queries about the data would first aggregate all of the data onto a central server before performing model optimization on all data simultaneously. In the FL setting though, all data remains with the individual client and only the statistical model resides on the central server. Optimization of the FL model occurs in a distributed fashion, where copies of the model are sent to clients and model *updates* are aggregated from clients to the central server at each optimization step. Here, individual updates are calculated only using the data on the client device.

Exact implementation details of FL depend on the statistical model being constructed, but its main benefit over the centralized regime is that a security breach of the central server is far less serious. At worst, the attacker can only access historical communications between the central server and clients, none of which will contain any private user data. However, FL can reduce the utility of the central model[9] and slow optimization depending on the availability of individual clients. Nevertheless, initial research on FL in a mHealth context has shown that models trained in a federated manner suffer only minor performance costs compared to models trained centrally[10]. In this regard, FL would seem like a prime candidate for privacy protection in large-scale mHealth applications, but we show that FL alone is not sufficient to defend against all privacy threats.

### Differential Privacy

Differential Privacy (DP) aims to overcome the problems associated with information leakage that occur whenever a query is conducted on a private dataset. In short, if the answer to a query (e.g. “What is the average age of patients in XYZ hospital?”) given a fixed dataset is deterministic, then a clever attacker could combine a series of queries with auxiliary information to infer private information about individual records in the dataset. This includes whether a particular record is in the dataset (membership inference) or features associated with a particular record (data reconstruction).

DP protects against such attacks by adding a stochastic component to query outputs. For example, when the output is a continuous value, this is often accomplished by adding some amount of random noise to the answer. Since there is no longer a deterministic mapping between dataset properties and query outputs, the probability of an attacker successfully inferring private information is reduced. In conventional “□-□ Differential Privacy,” we say that a mechanism *M* that generates an output based on an input dataset *D* is □-□ differentially private if the following inequality holds:

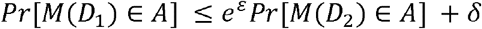

where *A* is any set of possible output values and *D*_*1*_, *D*_*2*_ differ only in the presence/absence of a single record[11]. In other words, a differentially private construction provides strong bounds on the probability of a successful membership inference attack by enforcing that the model output is sufficiently close (defined by epsilon and delta) in distribution to what the model would have output if one user’s record was deleted from the dataset.

As with FL, the exact implementation of DP depends on the nature of queries made on the private dataset. DP is commonly used to control information leakage that results from exposing parameters of models trained on private data. For example, Abadi, et al. demonstrate how DP can be integrated into gradient descent algorithms for deep learning to achieve certain statistical guarantees of privacy[12]. As with FL, the benefits of DP come at the cost of lower model utility, often embodied in lower model accuracy and higher optimization times.

### Prior Work

FL has found widespread applications in healthcare settings. Using electronic health records distributed across hospital systems, researchers have successfully developed federated models to predict heart-related hospitalizations[13], ECG classification[14], and clinical outcomes for COVID-19 patients[15]. In addition to the results in[10], other research has shown that FL can be applied to wearable sensor data in biomedical applications[16], a particular area of interest for mHealth research. Such studies generally find that the benefits of FL outweigh the costs.

Similarly, there have been many uses of DP in healthcare applications[17], including drug sensitivity prediction[18] and coronary heart disease diagnosis[19]. Other work has combined DP with FL to achieve enhanced privacy protection[20]. However, such studies have not demonstrated a universal rule for choosing the ε DP parameter in any given application. In fact, Hsu, et al. find that optimal choices of ε in existing literature span orders of magnitude depending on context[21]. This paper therefore hopes to contribute to the DP literature by evaluating its efficacy in a novel health sensor data application.

Although quantifying model utility is straightforward in most cases, quantifying privacy protection is usually an imprecise exercise. The benefits of FL are clear on a conceptual level (eliminating risks associated with centralizing data in one location), but how this translates to a quantifiable increase in privacy protection is unclear. While DP provides strong statistical guarantees tied to numeric parameters (such as in □-□ DP), such guarantees only apply to a specific class of privacy attacks (membership inference attacks). For instance, Naseri, et al. show that FL and DP may not protect against property inference attacks that aim to uncover private attributes of individual client datasets[22].

In this paper, we measure the privacy-utility tradeoff of FL and DP using wearable sensor data gathered from a large-scale clinical study of medical interns. By simulating a privacy attack against a model constructed using this dataset, we measure the privacy protection offered through FL and DP. To the best of our knowledge, this is the first paper to 1) evaluate the effectiveness of FL and DP on a real-world health sensor dataset, and 2) measure the privacy protection of FL and DP in the context of a simulated attack.

## Methods

We must construct models of a target system and an external attack to evaluate the strength of privacy protection mechanisms in guarding against actual privacy attacks. The **Target System** will be a prediction model existing on a central server that implements FL and DP in an attempt to protect against privacy threats. The **External Attack** will simulate an adversarial actor intent on uncovering private information about individuals in the Target System. Enacting the External Attack against the Target System will allow us to assess the performance of the Target System in terms of model utility and privacy protection.

### Data: Intern Health Study

Data for this paper come from the 2017-2019 cohorts of the University of Michigan Intern Health Study (IHS)[23]. IHS aims to investigate the biological and genetic factors affecting the relationship between stress and depression. The study follows medical interns at several dozen facilities in the United States and China. In addition to providing demographic information at the beginning of the study, participants are asked to wear a Fitbit device for the duration of their internship and complete daily mood EMAs (Ecological Momentary Assessments) through a mobile app.

The data originally covers 6,660 registered participants with 1,241,629 daily sensor observations. After data cleaning (see Table 1 for details), our final dataset contains 4,274 participants and 596,585 daily sensor observations. Although the IHS data includes information about participants’ medical internships (such as specialty), we exclude this information in order to better simulate a mHealth system that would be used with a general population. Pertinent demographic features of the data are given in Tables 1 and 2, while Table 3 shows summary statistics for participant age, daily mood, and daily sensor data. Table 4 summarizes the degree of missingness for each sensor measurement over all daily observations.

**Table 1:** Participants and Sensor Data by Cohort Year

| Cohort Year | Participants (Raw Data) | Participants (Clean Data*) | Daily Sensor Obs. (Raw Data) | Daily Sensor Obs. (Clean Data) |
| --- | --- | --- | --- | --- |
| 2017 | 2,846 | 531 | 119,850 | 83,999 |
| 2018 | 2,129 | 2,098 | 576,535 | 277,754 |
| 2019 | 1,685 | 1,645 | 539,158 | 234,832 |
| **Total** | 6,660 | 4,274 | **\*\*1,235,543** | 596,585 |

\* Participants with no available sensor data and sensor observations not linked to registered participants are removed. We also exclude 40 participants with invalid/missing age values, 2 participants with missing sex values, and 26 participants with missing ethnicity values. Sensor observations with no mood score are also removed.
\*\* This number is slightly less than the original dataset size due to observations not linked to registered participants.

**Table 2:** Breakdown of Participants by Ethnicity and Gender (Clean Data)

|  | Male | Female |
| --- | --- | --- |
| **White** | 1,072 | 1,354 |
| **Black / African American** | 73 | 148 |
| **Latino / Hispanic** | 87 | 81 |
| **Asian (e.g. Indian, Chinese)** | 456 | 524 |
| **Native American** | 1 | 3 |
| **Pacific Islander** | 1 | 0 |
| **Other** | 10 | 8 |
| **Multi-racial** | 156 | 223 |
| **Arab / Middle Eastern** | 40 | 37 |
| **Total** | 1,896 | 2,378 |

**Table 3:** Descriptive Statistics of Daily Sensor Data and Participant Age (Clean Data)

|  | **Mean** | **Median** | **Std Dev** | **Min** | **Max** |
| --- | --- | --- | --- | --- | --- |
| **Participant Age (Years)** | 27.66 | 27 | 2.62 | 17 | 47 |
| **Mood EMA Score** | 7.27 | 7 | 1.63 | 1 | 10 |
| **In-bed Minutes** | 417.98 | 437 | 137.59 | 4 | 1379 |
| **Sleep Minutes** | 372.11 | 389 | 123.81 | 0 | 1228 |
| **Active Minutes** | 63.83 | 25 | 92.6 | 0 | 1016 |
| **Step Count** | 8,819 | 8,215 | 4,739 | 1 | 70,138 |
| **Resting Heart Rate (BPM)** | 62.83 | 63 | 7.63 | 39 | 100 |

**Table 4:** Daily Sensor Data Missingness (Clean Data)

|  | **% Days Missing\*** | **Avg Days of Data per Participant** |
| --- | --- | --- |
| **In-bed Minutes** | 43.52% | 78.84 |
| **Sleep Minutes** | 43.52% | 78.84 |
| **Active Minutes** | 58.87% | 57.42 |
| **Step Count** | 31.21% | 96.02 |
| **Resting Heart Rate** | 54.26% | 63.84 |
| **Combined Features\*\*** | 71.86% | 39.28 |

\* Missingness is measured relative to the baseline obtained after removing all days with missing mood score and all participants with missing demographic features or no sensor data.
\*\* A non-missing day for this row is any day with all five sensor measurements recorded.

### Data Preprocessing

The most significant step in preparing the IHS data for analysis is dealing with missing data. Even though a typical medical internship lasts 12 months, the average participant in the study logged a bit more than one (not necessarily continuous) month of complete sensor data. Given our objective of building a real-world mHealth system, we cannot simply exclude all observations with any missing feature. Furthermore, exploratory tests demonstrate that neural network classifiers trained on only complete cases are not useful predictors of mood score. Therefore, we impute missing sensor features using 10 iterations of MICE due to its flexibility and relatively low computational cost^24^. We acknowledge that true FL would require us to impute data locally at the participant level, but data availability considerations lead us to adopt a centralized imputation approach.

Our feature set includes the 9 features (Cohort Year, Age, Sex, Ethnicity, and 5 sensor measurements) listed in the previous section, as well as one-day and two-day lags for each of the 5 sensor measurements and the mood score. Lags account for the possibility that certain “sensor events” (such as one night of poor sleep) may have a delayed impact on mood. Additionally, we include 3 time-based features indicating the observation’s day of week, day of month, and day of year to account for unmeasured factors that may be correlated with time. Lastly, we perform common preprocessing steps to our feature set, such as standardizing continuous features and splitting categorical features into component binary features.

### Target System Construction

In the Target System, the statistical models live on the central server and use data from individual IHS participant devices for training. We formulate both a regression and classification task for the Target System and train models for each using our sensor and demographic data. These two tasks provide robustness in assessing the effectiveness of our privacy protection measures. The regression task predicts mood score on the 1-10 scale, while the binary classification task predicts whether the user’s mood has improved from the previous day. Since we do not consider model interpretability in our analysis, we implement neural networks for both tasks, owing to their flexibility and accuracy advantages over other machine learning methods. Details of our implementation are given in **Appendix I**. This paper reports results from the regression task and leaves results from the binary prediction task in the attached **Appendix II**.

### External Attack: Underlying Assumptions

A multitude of methods could be designed to compromise user privacy through the Target System. For example, attackers interested in whether a specific user/record appears in the Target System may mount a *membership inference attack*, whereas those interested in ascertaining certain statistical properties of the private training data (either globally or on a per-user basis) may mount a *property inference attack*. Other attackers may attempt *data reconstruction attacks*, which aim at reconstructing partial or whole records from the original training data^22^. Our attack model derives from domain-specific considerations regarding the environment of mHealth applications, the privacy demands of system users, and the identities of possible attackers.

In mHealth systems containing a large portion of the general population, membership inference has limited value and reconstruction of a specific user’s data is quite difficult. Therefore, we assume the attacker is interested in **property inference** on individual user data, particularly whether an IHS participant has an average daily mood EMA score higher than the global average daily mood EMA score (denoted hereafter by “mood status”). Such inference is relatively easy to execute and could expose sensitive information about the participants; for instance, those with consistently low mood scores may be at higher risk of depression or other mental disorders. Regardless of the attacker’s exact purposes, the mere possibility of a successful inference of mood status may significantly undermine public trust in the mHealth system.

The literature on privacy attacks provides two broad dimensions along which a privacy threat may be assessed based on the attacker’s resources. The first dimension is the attacker’s level of access to the Target System and any associated privacy protection systems. The literature often differentiates between “black box” model access, where the attacker is limited to viewing only the Target System’s model output for a given input, and “white box” access, where the attacker is able to view the model architecture and all related parameters along with the details of any privacy protection mechanisms. The second dimension is the attacker’s ability to alter model parameters to their liking. Certain attacks can be implemented passively, meaning the attacker can compromise user privacy by simply observing changes to the Target System’s statistical model that occur during training. Others require the attacker to actively influence model parameters, usually by injecting their own customized training data into the system.

We assume the attacker has **white-box access** to the Target System’s central server (including the statistical model and all communications with the server) but can only carry out privacy attacks **passively**. We also assume the attacker has no ability to access any “live” individual user devices in the Target System (any devices actively participating in model training). These assumptions match the “rogue employee” profile, an insider who is easily able to access confidential details about the Target System, but would not be able to effect changes in model parameters without raising suspicion. This profile was selected to balance plausibility with preparation for a worst-case scenario. While it is unlikely that any adversarial actor could influence the training process of the Target System, we believe any serious actor would likely gain insider access somehow.

### External Attack: Implementation

The implementation details of this attack follow[25]. The attack is carried out in the three stages described below. For this analysis, we assume the statistical model in the Target System is trained using FL and Local DP, implying individual user devices do not share their data and add noise to their gradient updates before sending them to the central server. Additional details can be found in the next section “Implementing Federated Learning and Differential Privacy.”

#### Stage 1: Accessing Target Model Parameters

Given the attacker has full access to all information stored on the central server at training time, we assume they observe Target System model parameters at time *t* (given by θ_*t*_) as well as the gradient updates (given by ∇_i, t_) with respect to θ_*t*_ sent to the server from individual user devices *A*_*i*_. The relationship between these variables is shown on the left side of Figure 1.

**Figure 1:**
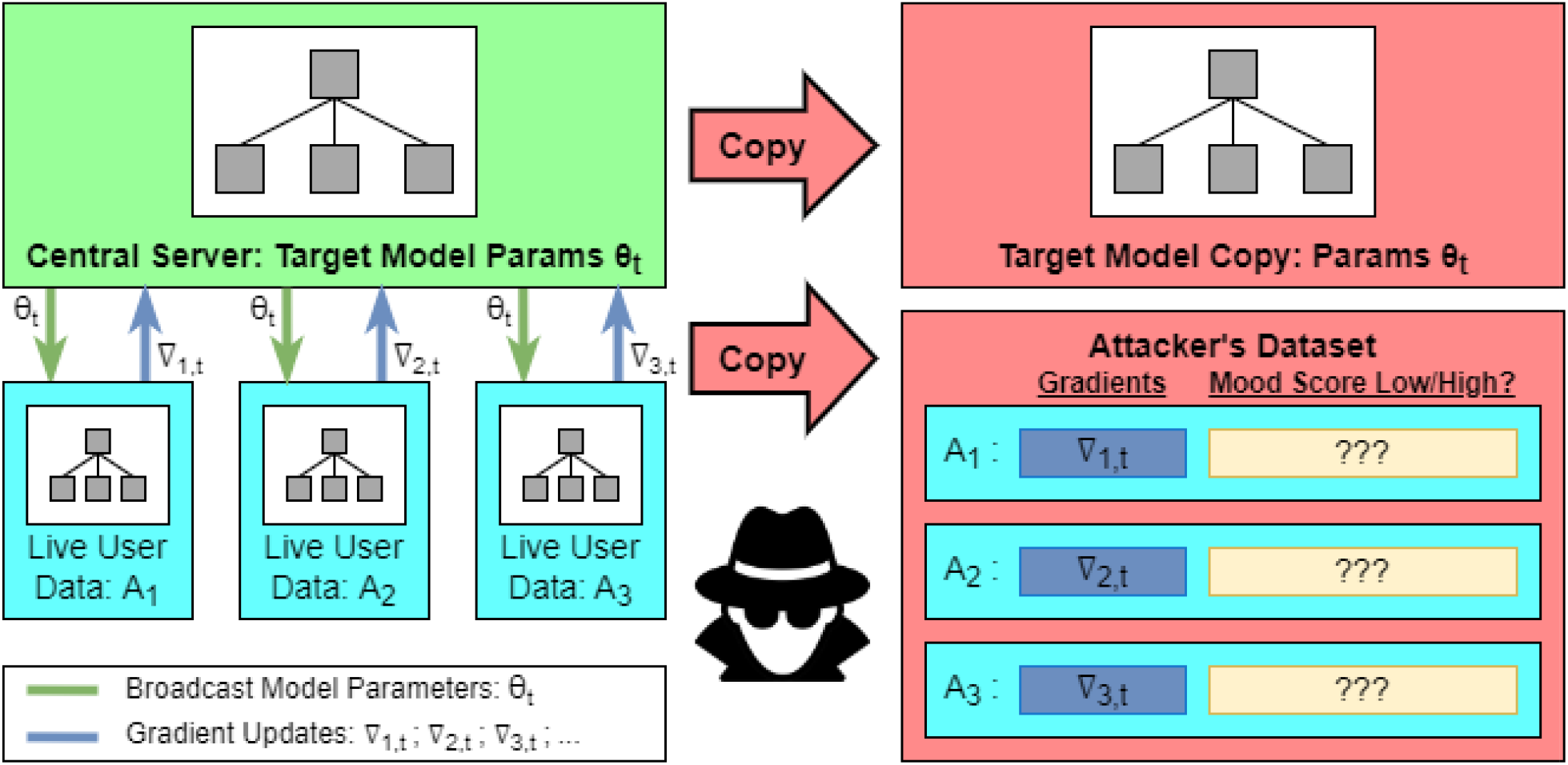
Attacker Infiltration of Central Server

The attacker first copies the entire Target Model (including parameters θ_*t*_) as well as the gradient updates ∇_i, t_. The attacker will ultimately attempt to predict the mood status of each user *A*_*i*_. Note that under DP, the gradient updates ∇_i, t_ observed by the attacker would include any noise added to the true model gradient on the user device.

#### Stage 2: Constructing the Attacker’s Dataset

In order to predict mood status for live users, the attacker requires an auxiliary collection of users for whom mood status is known and model gradients with respect to θ_*t*_ can be calculated. We assume the attacker can access such a dataset because they already have access to the central server. This data could be sourced from central server databases containing raw data for pilot users, a small number of previously compromised user devices, an external published dataset, or even users working in collusion with the attacker. The process of using this data to construct the attacker’s dataset is shown in Figure 2. In particular, each user in the auxiliary data has known mood status and generates a gradient ∇’_i, t_ with respect to the same Target Model parameters θ_*t*_. This gradient contains the same amount of noise as the observed gradient updates from live users since we assume that the parameters used for privacy protection in the

**Figure 2:**
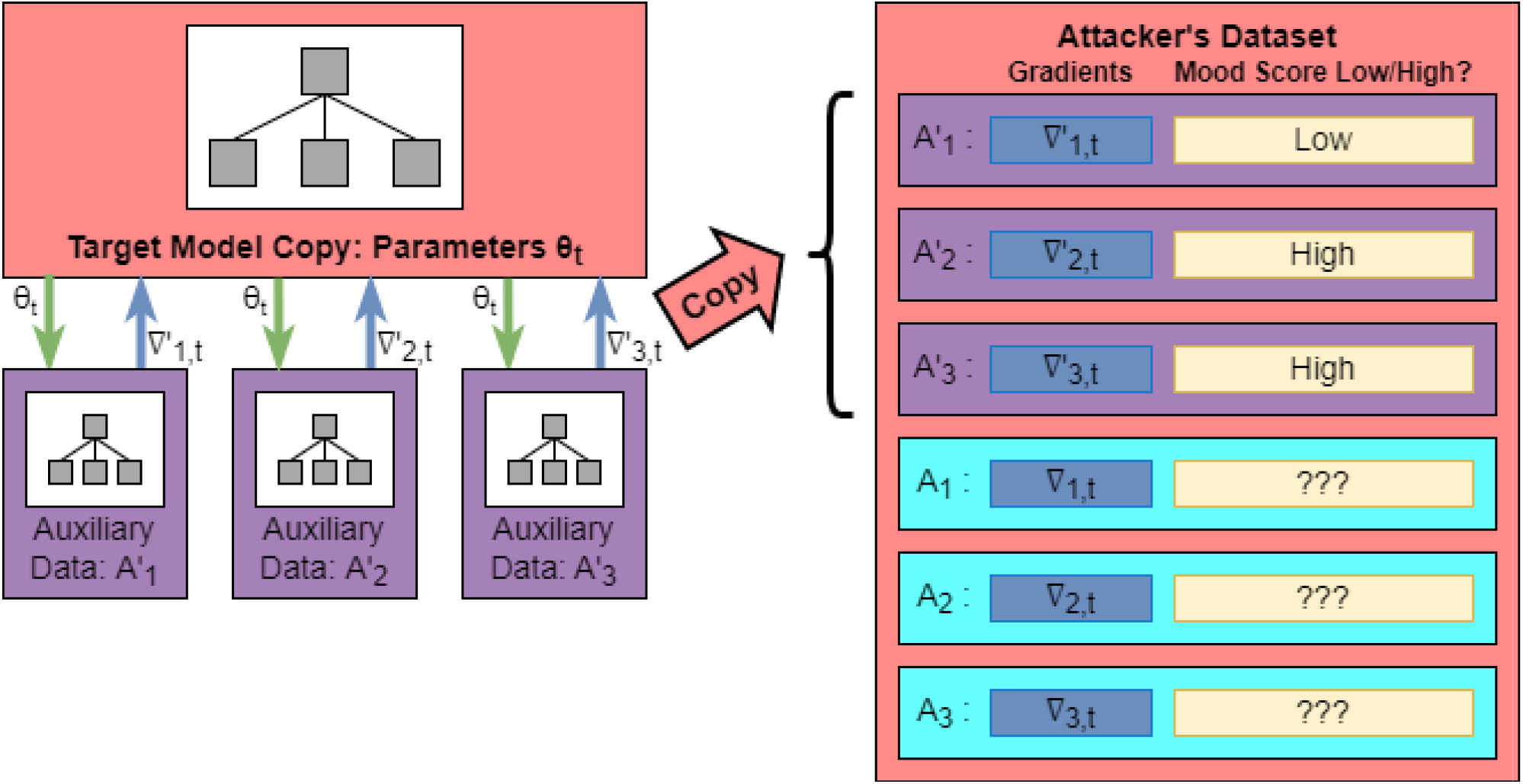
Constructing the Attacker’s Dataset

Target System are stored on the central server. We reason this assumption is feasible for systems implementing FL and DP because they cannot broadcast the relevant parameters to all users while hiding them from adversaries with access to the central server. This increases the likelihood of successful inference relative to if the noise parameter was not known.

#### Stage 3: Mood Status Prediction

Finally, the records in the attacker’s dataset for which mood status is observed can be used to train a binary “batch property classifier” that predicts mood status (labels) from the provided gradients (features). This process is visualized in Figure 3. Any machine learning model can be used in this step, but we adopt a neural network approach following the example of [25]. The details of our implementation are given in **Appendix I**.

**Figure 3:**
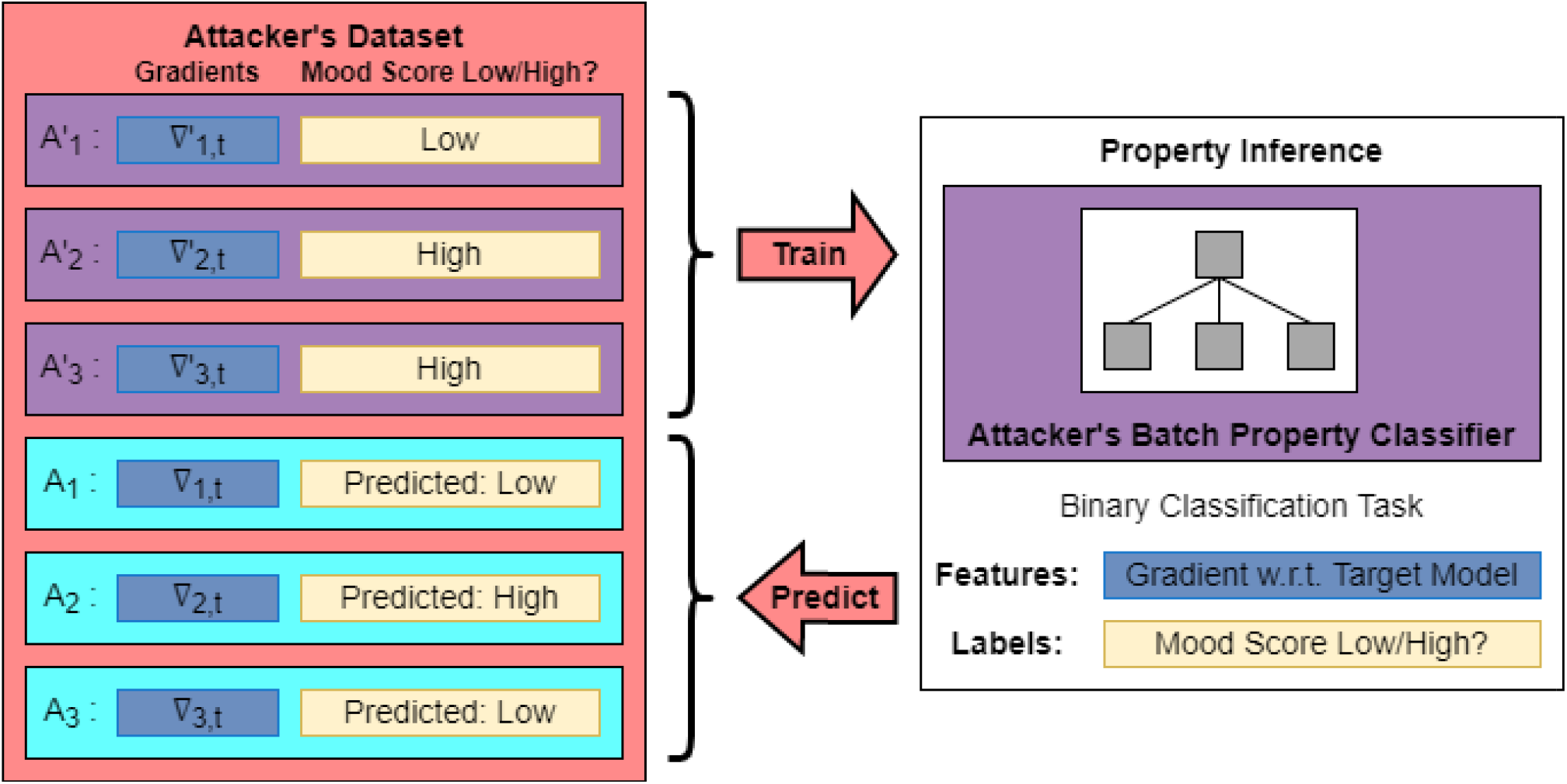
External Attack Training and Final Inference

### Implementing Federated Learning and Differential Privacy

Our proposed defense against the External Attack is using local Differential Privacy (DP) techniques in conjunction with Federated Learning (FL) to mask the information present in the gradient updates communicated to the central server. The specific implementation follows [22]. The general algorithm is shown in Figure 4.

**Figure 4:**
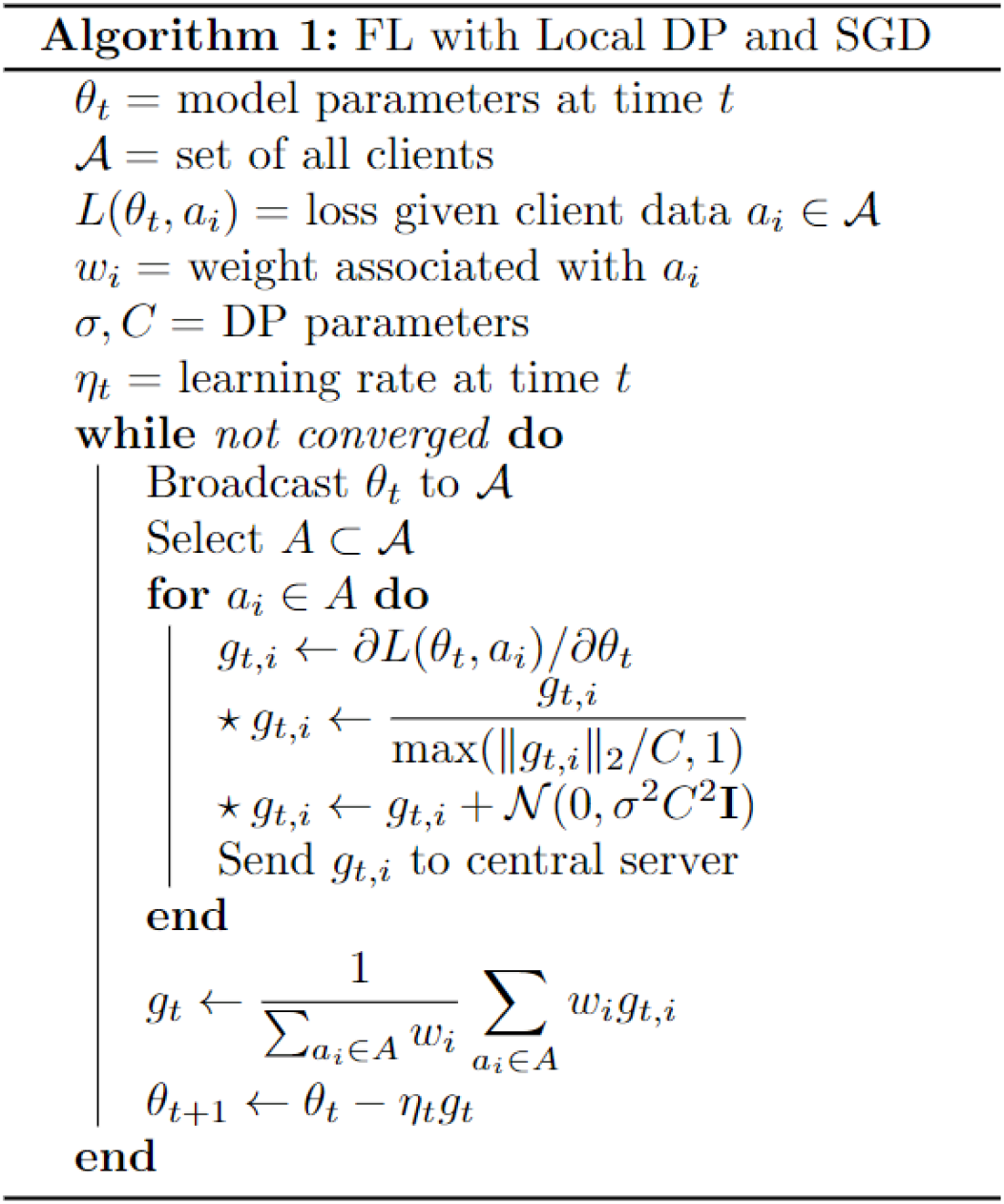
Pseudocode for Privacy Protection Implementation

Our implementation of FL randomly samples 30% of the study participants in each iteration to participate in the corresponding model update (set *A*). Our procedure is very similar to vanilla FL and differs only in the second and third steps (indicated with a star) of the inner for loop, which are described below:

1. Clip each computed gradient to have *L*_*2*_ norm of at most *C*
2. Add gaussian noise with mean zero and variance σ^*2*^*C*^*2*^ to each gradient component independently.

The first step ensures that the signals within the gradient updates do not overpower the noise, while the second step provides the stochastic component that addresses information leakage resulting from deterministic gradient calculations. For this analysis, we set *C = 1* for all cases and vary the σ parameter to achieve different levels of privacy protection. Note that each choice of σ and *C* corresponds to a particular level of ε in the traditional DP definition. We choose the former parameterization for ease of interpretation, especially since ε is mainly formulated in the context of membership inference attacks. Other parameters remain identical to those outlined in **Appendix I** for the Target System.

### Performance vs Privacy Protection: A Simulation Approach

In general, it is difficult to quantify the effectiveness of the External Attack against the Target System, so we utilize a simulation-based approach to report results for several different attacker settings. We vary σ, the noise parameter in constructing the stochastic gradients, to test different levels of privacy protection. We also model differences in the attacker’s access to auxiliary user data by changing *q*, the proportion of IHS study participants with “compromised devices” whose data is available to the attacker. Figure 5 shows the basic simulation setup for our analyses.

**Figure 5:**
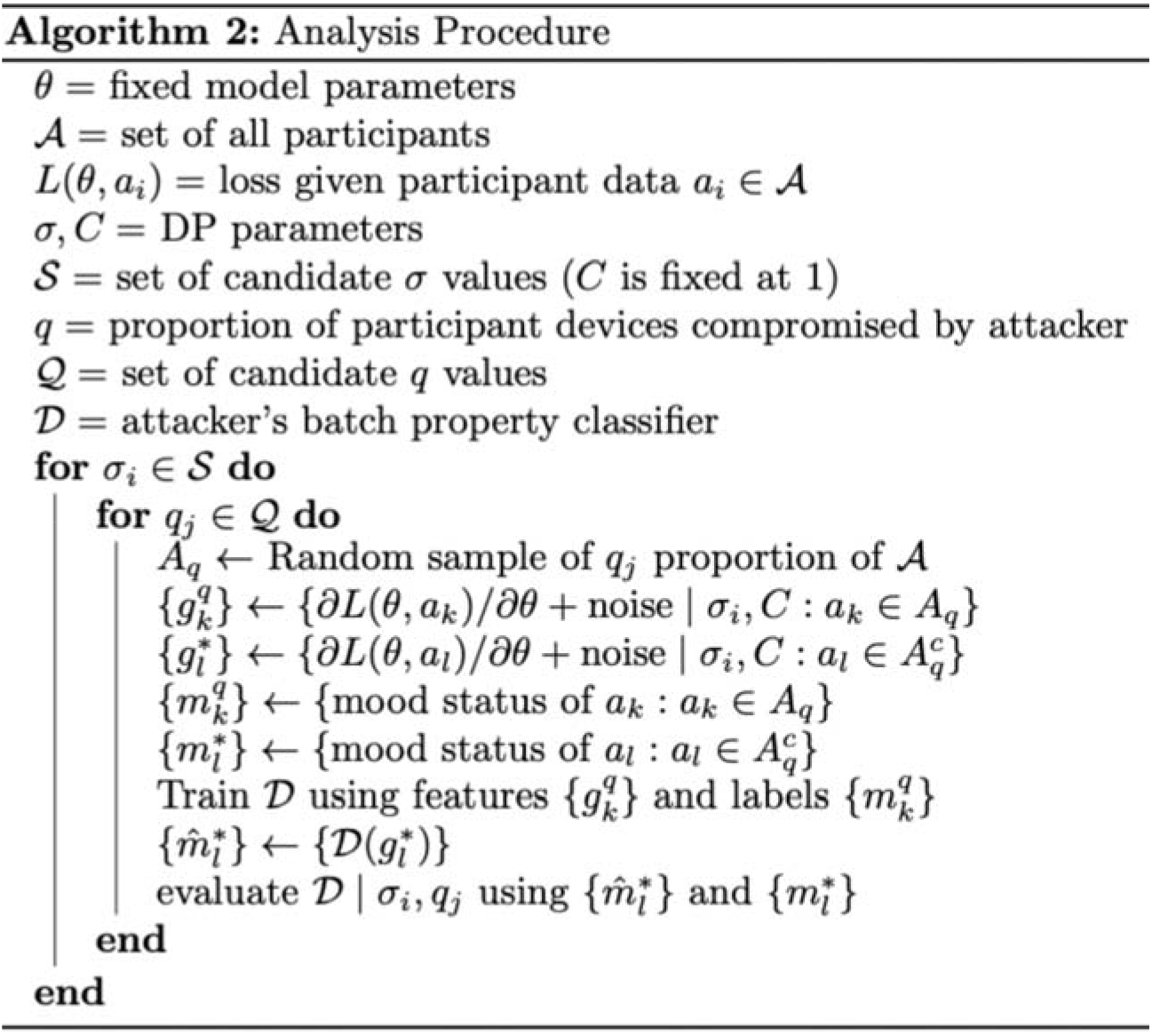
Pseudocode for Our Assessment of External Attack Effectiveness

#### Measuring Costs of Privacy Protection

Higher levels of privacy protection generally hinder the Target System’s ability to train the model effectively (since the gradients have more added noise). We measure this cost in terms of training time and final model utility for different values of σ. To facilitate comparability, we initialize the Target Model identically for each case and seed all stochastic components in the training process (selection of clients, network dropout, etc) identically.

#### Measuring Effectiveness of Privacy Protection

We measure the effectiveness of our privacy protection procedure by the External Attack’s ability to infer the mood status of non-compromised users. Designating a fixed subset of such users as a “test set”, we record the maximum test AUC achieved by the attacker’s classifier *at any point during its training process (up to 600 epochs)*. Although the attacker theoretically does not have access to testing labels, this procedure gives an experimental upper bound on their performance. We also calculate model sensitivity and PPV as more easily interpretable metrics. All metrics are calculated for each choice of σ and *q*. To facilitate comparability, all evaluations are performed with the attacker using the same Target Model parameters produced after 20 training epochs to generate their gradient dataset. All other stochastic components are seeded identically for each evaluation.

### Data Availability Statement

The data that support the findings of this study are available on reasonable request from the corresponding author Alexander Shen. The data are not publicly available due to considerations regarding participant privacy.

### Code Availability Statement

The computer code that supports the findings of this study are available on reasonable request from the corresponding author Alexander Shen. The code is not publicly available due to considerations regarding participant privacy. See the **Reproducibility** section for a version of the code developed for publicly available data.

## Results

### Target Model Training Metrics

Figure 6 plots the Target System’s model loss and accuracy metrics over each gradient update. Training progress under the conventional centralized training protocol is also plotted for reference. A σ value of zero denotes FL with no DP implementation. We see a clear effect of both FL and DP on the training process for the Target System’s statistical model. Relative to the centralized protocol, FL leads to a noticeable increase in training time, but the final model accuracy seems to be very similar (at least within the given training window). The addition of noise via DP seems to substantially affect both training time and final model accuracy. Interestingly, training loss seems to begin increasing at some point in the training process when additional noise is added, potentially indicating shortcomings in conventional optimization techniques when using both FL and DP. Figure 7 shows the relationship between training time/final model utility and noise scale, where training time is defined as the number of gradient updates needed for model loss to fall within 1% of the minimum loss over 1000 updates.

**Figure 6:**
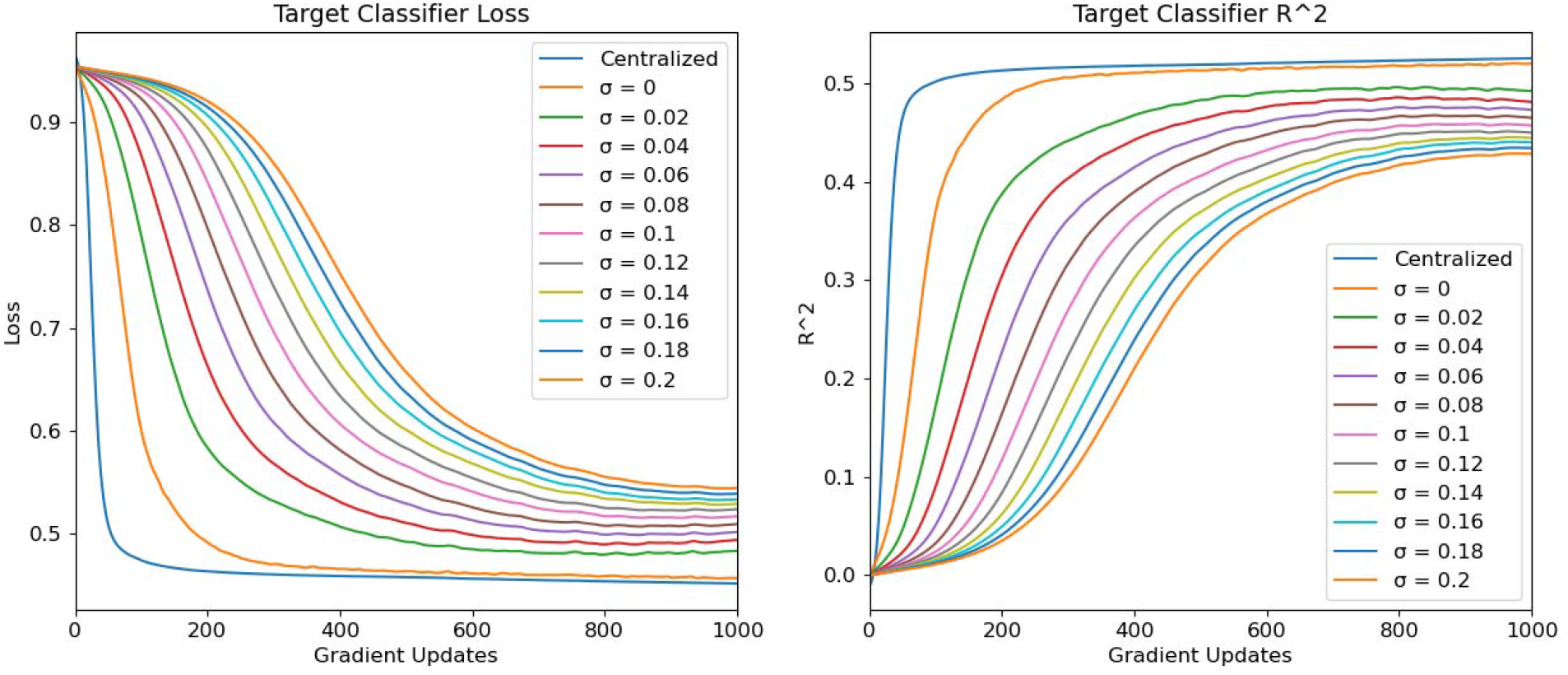
Target System Model Performance In all simulations, training runs for at most 1000 epochs

**Figure 7:**
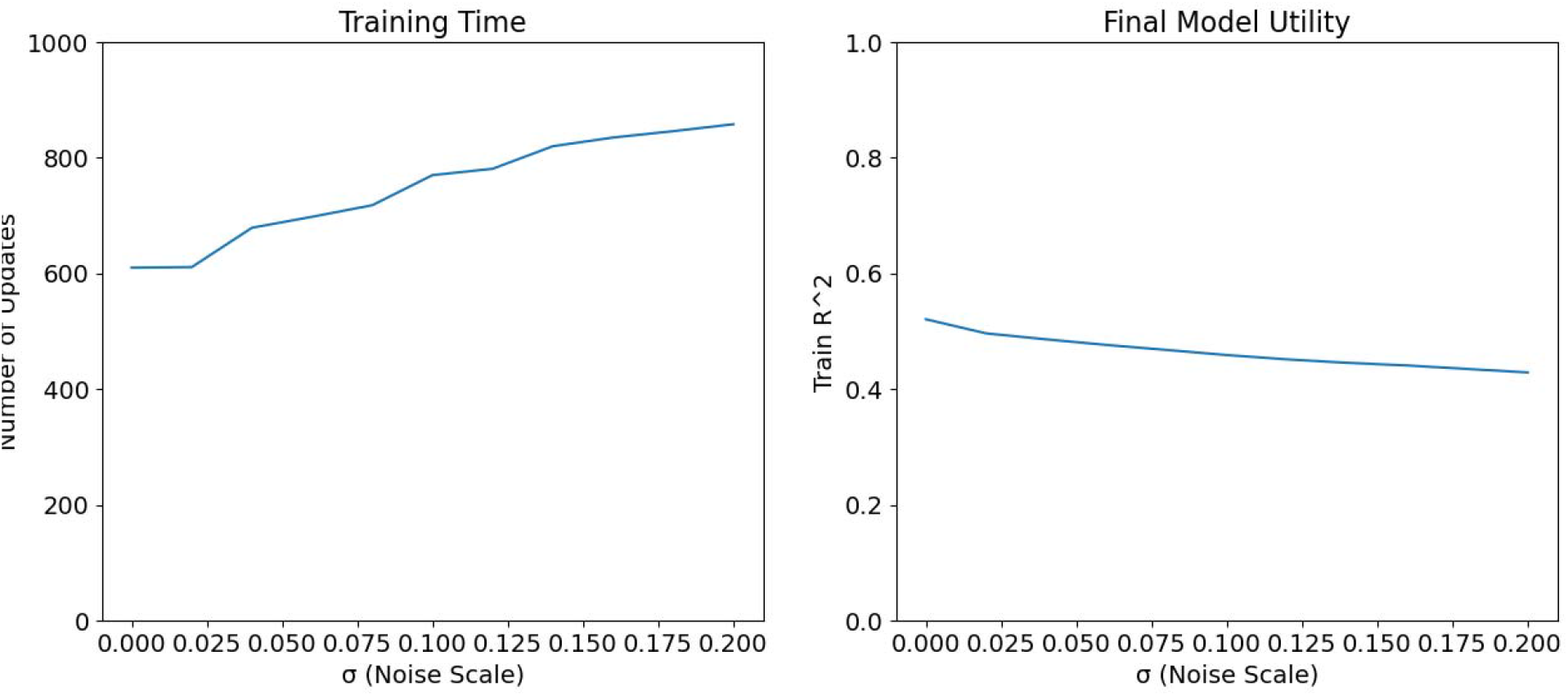
Training Time/Final Model Utility vs. Noise Scale

### Privacy Protection Metrics

Figure 8 shows the External Attack’s success in inferring a participant’s mood status based on the participant’s observed gradient to the fixed Target System model parameters. Immediately, we note that FL alone (zero noise) is insufficient for protecting against this type of attack. Even if the attacker has access to labeled data for only 10 participants (out of 4,274 total participants), they can successfully infer the mood status of a large majority of all other participants. However, adding noise to the gradient updates significantly decreases the attacker’s performance, even when they have access to a large amount of labeled training data. Figure 9 shows similar results for the attack’s positive predictive value and sensitivity, given that a low mood status is viewed as a “positive test result.”

**Figure 8:**
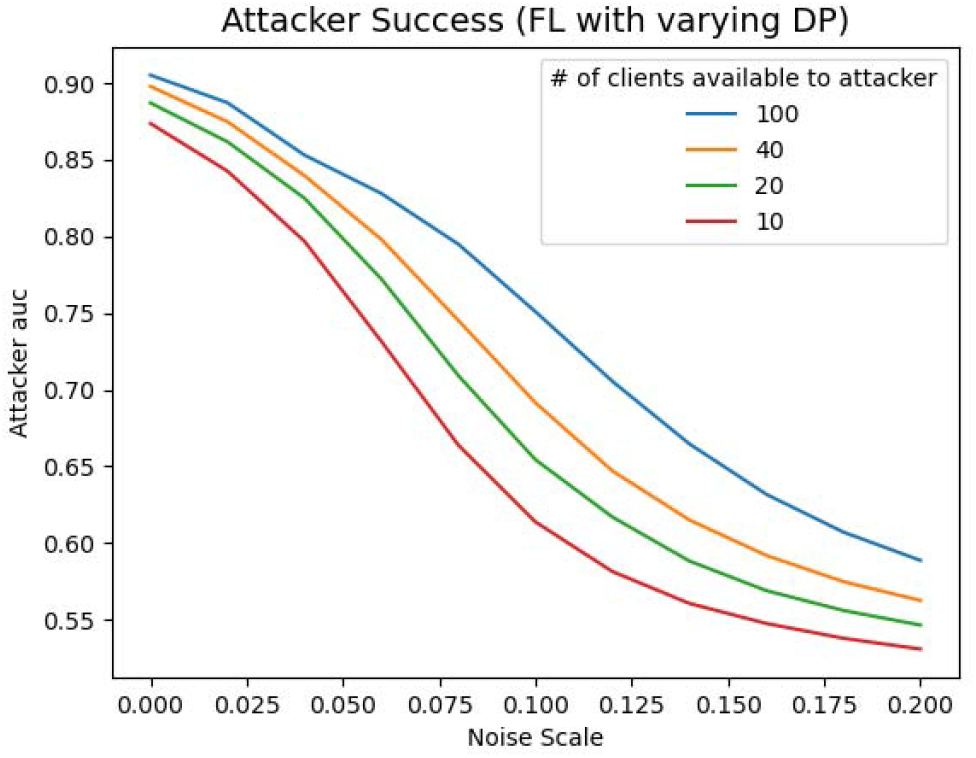
Attacker Success vs. Noise Scale

**Figure 9:**
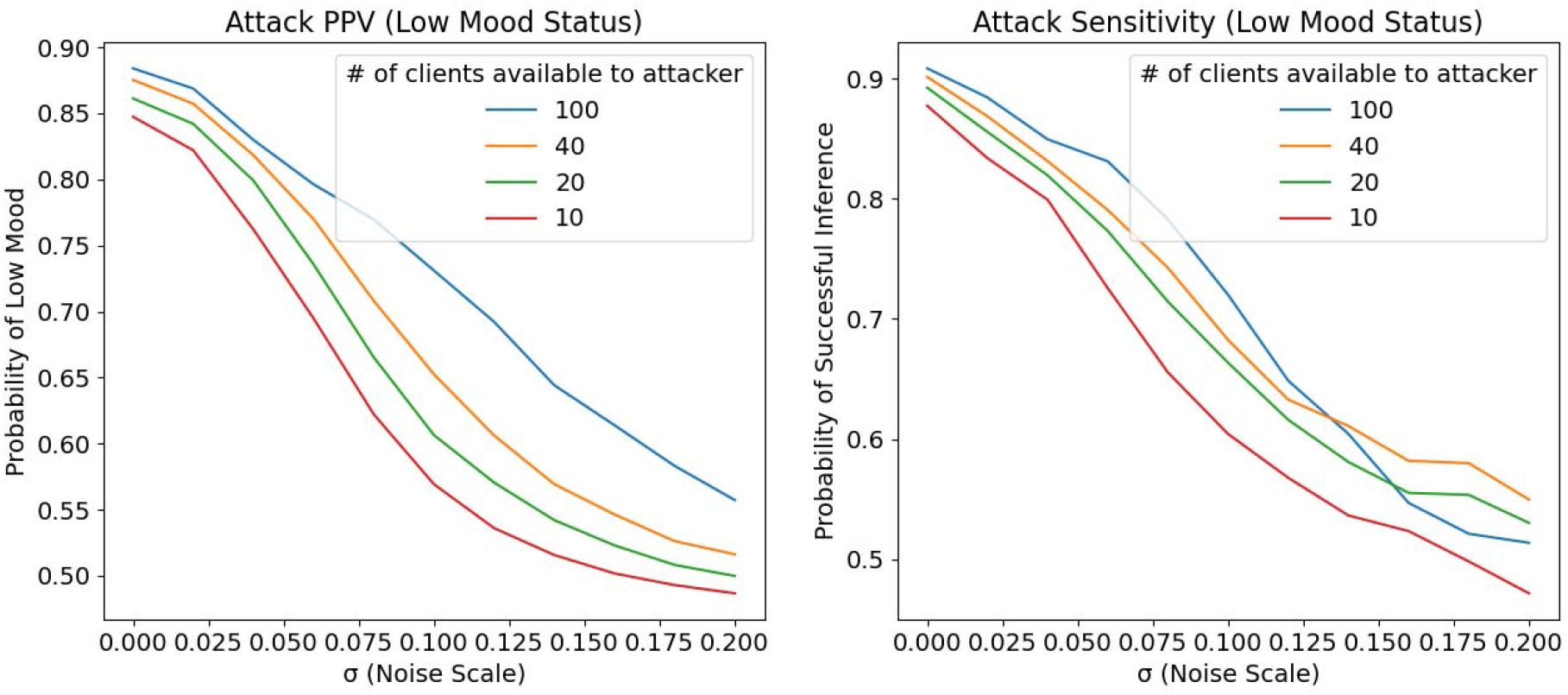
Attack PPV/Sensitivity vs. Noise Scale

Combined with the training metrics from the previous section, we can start to assign concrete tradeoffs between model utility and privacy protection in the context of this particular Target Model. For example, if the attacker has access to 100 participants’ labeled training data, setting the noise scale to 0.1 increases training time by 26.2%, decreases final model R^2^ by 11.5%, and decreases the privacy attack’s PPV by 17% relative to FL with no additional noise. Increasing the noise scale to 0.2 increases training time by 40.7%, decreases final model R^2^ by 17.3%, and decreases the privacy attack’s PPV by 36.4% compared to FL with no noise.

### Vulnerability Metrics

This section analyzes the benefits of our privacy protection procedure for various subgroups within our data. While we should strive to maximize privacy protection within reason for all members of the population, it is instructive to examine who is most at risk from this privacy threat and hence, who benefits the most from these privacy protection mechanisms.

Figure 10 shows the correlation between the attacker’s prediction of a particular participant’s mood status versus the participant’s actual mood status. The y-axis shows the attacker’s predicted probability of each participant having a high mood status, where values above 0.5 indicate participants ultimately classified as high mood status. As expected, there is strong positive association between the attacker’s prediction and the participant’s actual average mood when no additional noise is added to FL. This indicates that participants with average mood scores much lower than the global average are simultaneously at a much higher risk of a successful inference attack. Since privacy concerns are usually greatest for those with sensitive health conditions, this further underscores the insufficiency of FL alone. Fortunately, the addition of sufficient noise can eliminate this correlation, neutralizing privacy risks regardless of the participant’s mood tendencies. Similar analyses of privacy risk across gender, ethnicity, and age groups did not yield significant findings; the associated figures can be found in **Appendix I**.

**Figure 10:**
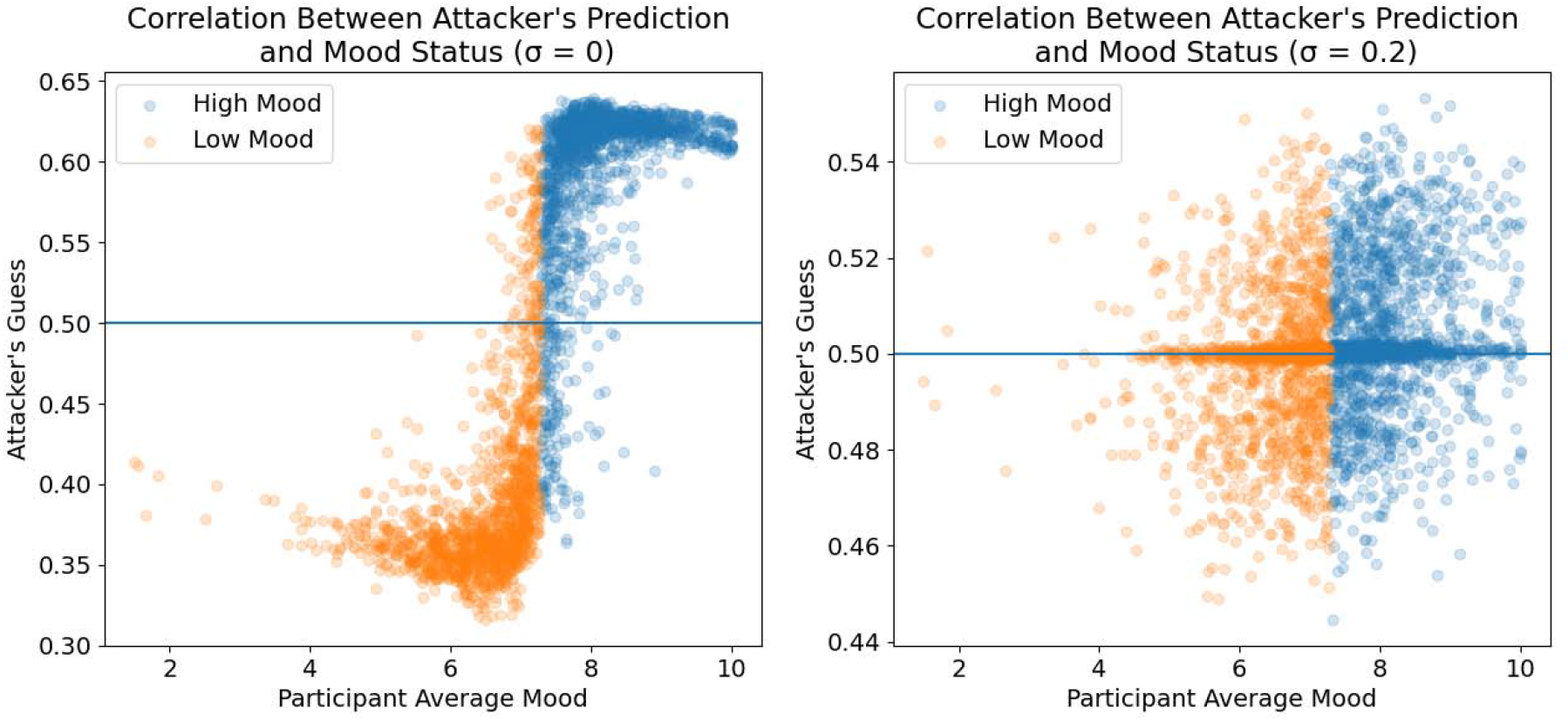
Attacker’s Predicted Mood Status vs. Participant Average Mood for Different DP Noise Parameters

## Conclusions

As mHealth applications continue arriving in the marketplace for the general public, our analysis provides important insight into future challenges of user privacy protection. We paint a clear picture of what the implementation of privacy protection technologies could look like in a real-world mHealth setting. Robust privacy protection necessarily comes at the cost of statistical utility, and our results illuminate the nature of that tradeoff using concrete metrics.

Our analysis also addresses a particular challenge in privacy research, namely the quantification and communication of the protection level offered by different methodologies. By narrowing the scope of the External Attack and adopting a simulation-based approach, we produce metrics that are both meaningful and easy to interpret. Through this process, we underscore two important principles for future work on privacy protection. First, our results show that FL, despite its simplicity and broad acceptance, is insufficient to protect against advanced privacy attacks on its own, emphasizing the need to evaluate and improve our existing tools for privacy protection. Our results also imply that mHealth users whose health indicators deviate strongly from the general population (i.e. those who most require privacy protection) are at higher risk under our threat model. This suggests the need for nuanced, context-specific approaches to privacy protection that consider the individual needs and vulnerabilities of system users.

While the results of this analysis are compelling, there are certainly many areas of improvement that could serve as the basis for further research. Our missing data imputation procedure is one of the main threats to the validity of our results, and it seems very likely that future mHealth studies will suffer from similar problems. Although our analysis does not devote attention to a rigorous solution to this problem, we acknowledge that imputation methods deserve the same scrutiny for privacy risks as the rest of the tools used for mHealth research. Additionally, our particular implementation of FL with local DP may not be highly optimized for our particular dataset, resulting in model metrics that overstate the cost of privacy protection. On the flip side, our implementation of the property inference attack may not exploit enough weaknesses in our Target System setup, resulting in an overstatement of the efficacy of our privacy protection measures.

More importantly, our simulation-based approach to measuring privacy protection sacrifices some statistical rigor in favor of interpretability. Unlike with most DP implementations, we cannot guarantee maximum bounds on the attacker’s probability of successful inference. It is not entirely clear how the mechanics of this attack change based on the Target System’s model architecture (neural network or otherwise), Target System’s classification task, attacker’s resources, attacker’s intended inference task, or properties of the underlying population. Still, we hope this work provides the foundation for the future development of numerical methods to approximate protection levels over a broader range of models and populations, or even theoretical bounds on the likelihood of successful property inference in mHealth systems.

It is our sincere hope that mHealth research continues to generate robust tools for privacy protection along with novel statistical methodologies and technical improvements. The weaknesses of FL clearly demonstrate the dangers of complacency; threats will continue to evolve, and our privacy protection technologies cannot fall behind. As mHealth applications continue to scale, safeguarding public trust must remain a top priority for researchers and practitioners alike.

## Data Availability

All data produced in the present study are available upon reasonable request to the authors

## Acknowledgments

Alexander Shen planned and performed the analysis and produced the results presented in this manuscript. He also performed background research necessary for simulating the privacy attack and implementing both Federated Learning and Differential Privacy on the Target System.

Alexander Shen is currently affiliated with Carnegie Mellon University, but the research described in this paper was performed during his time at University of Michigan.

Luke Francisco assisted in tasks for completing the final manuscript, including reviewing/proofreading, compiling references, and ensuring proper labeling of tables and figures.

Ambuj Tewari provided the initial idea for this analysis and pointed the team to relevant resources. Ambuj additionally provided frequent and impactful guidance during the course of the analysis, including reviews of the final manuscript.

Srijan Sen is the principal investigator for the University of Michigan Intern Health Study and provided the data used in this analysis. He also provided input on the final manuscript.

## Reproducibility

A version of the code used in this research was developed for use with the publicly available WESAD data. The code and instructions for downloading the WESAD data and running experiments is available at [26].

## Appendix I Additional Details/Results for the Regression Task

For the regression task in the Target System, we use a multilayer perceptron architecture for our neural networks with 256 nodes on the first hidden layer, 128 nodes on the second hidden layer, and 64 nodes on the third hidden layer. Leaky ReLU activation with slope 0.01 and dropout of 0.2 is applied between layers, except before the output layer. The networks are trained using the Adam optimizer with learning rate 0.0001, batch size 256, and Mean Squared Error (MSE) loss. Hyperparameters are tuned using 5-fold cross validation.

For the mood status prediction in the External Attack, we use a neural network classifier with 24 nodes in the first hidden layer, 20 nodes in the second hidden layer, and 16 nodes in the last hidden layer. Batchnorm, Leaky ReLU activation with slope 0.01, and dropout of 0.2 is implemented between layers, except before the output layer. The model is optimized using the Adam optimizer with learning rate 0.001 and weight decay 0.15 (since there are a large number of components in gradient updates to the Target Model). Hyperparameters are optimized using 5-fold cross validation.

Figure 11 shows the External Attack’s sensitivity when the attacker has access to data for 100 IHS participants, split by mood status and age. There seem to be differences across age, although overall trends are hard to identify.

**Figure 11:**
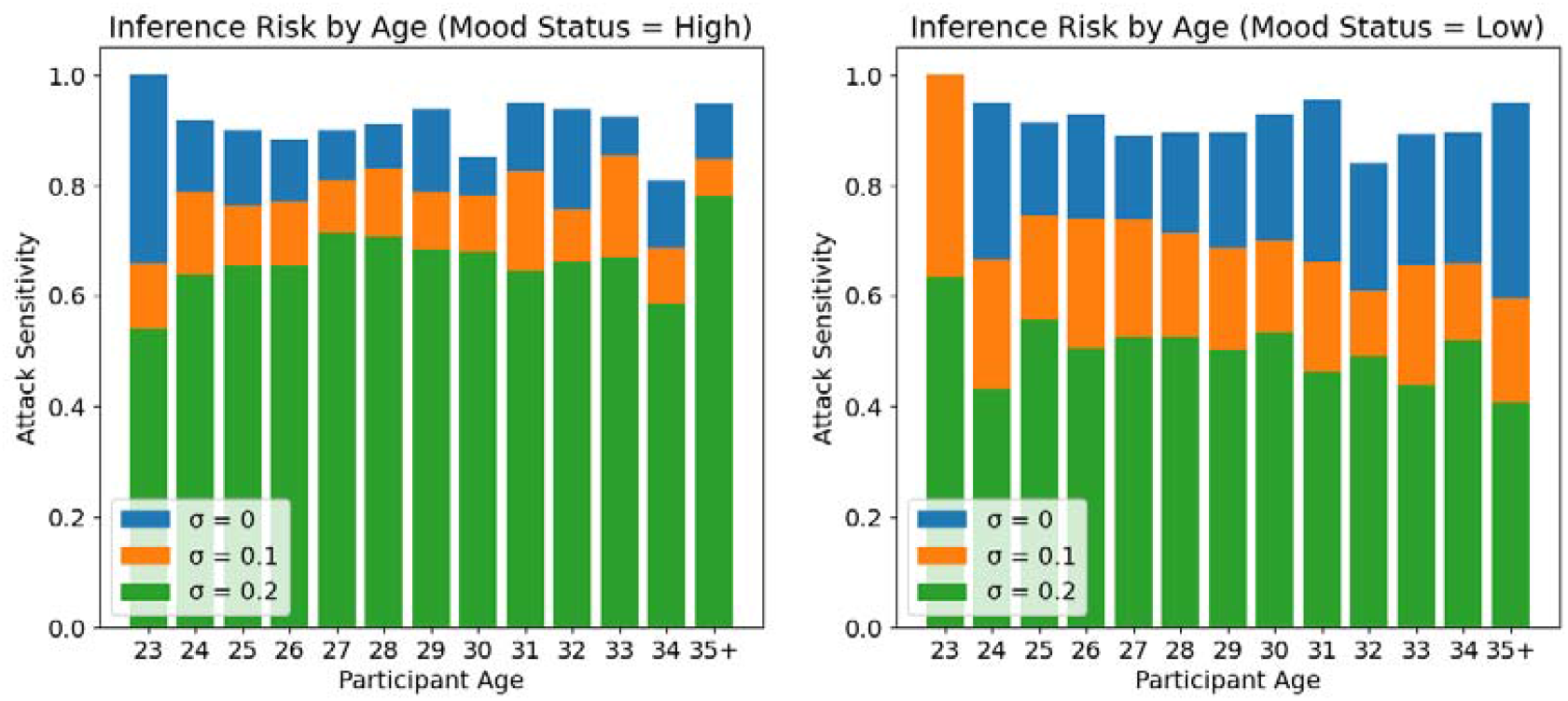
Attack Sensitivity by Age and Mood Status

Figure 12 shows the same data split by participant sex. There seem to be no significant differences between male and female participants.

**Figure 12:**
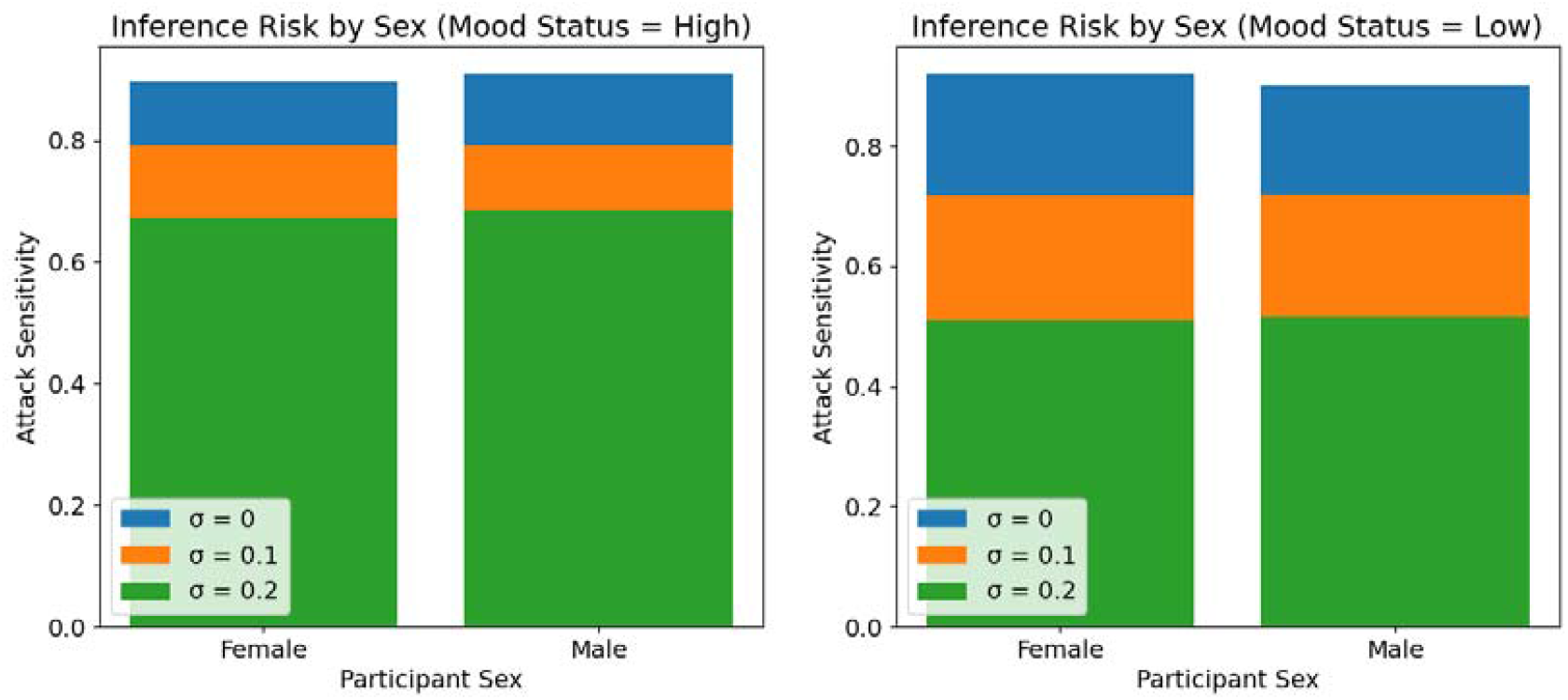
Attack Sensitivity by Sex and Mood Status

Figure 13 shows the same data split by participant ethnicity. For those with low mood status, adding additional noise to the Target System gradient updates seems to have heterogeneous effects on attack sensitivity, although small participant counts for some ethnic groups complicate drawing conclusions from these results.

**Figure 13:**
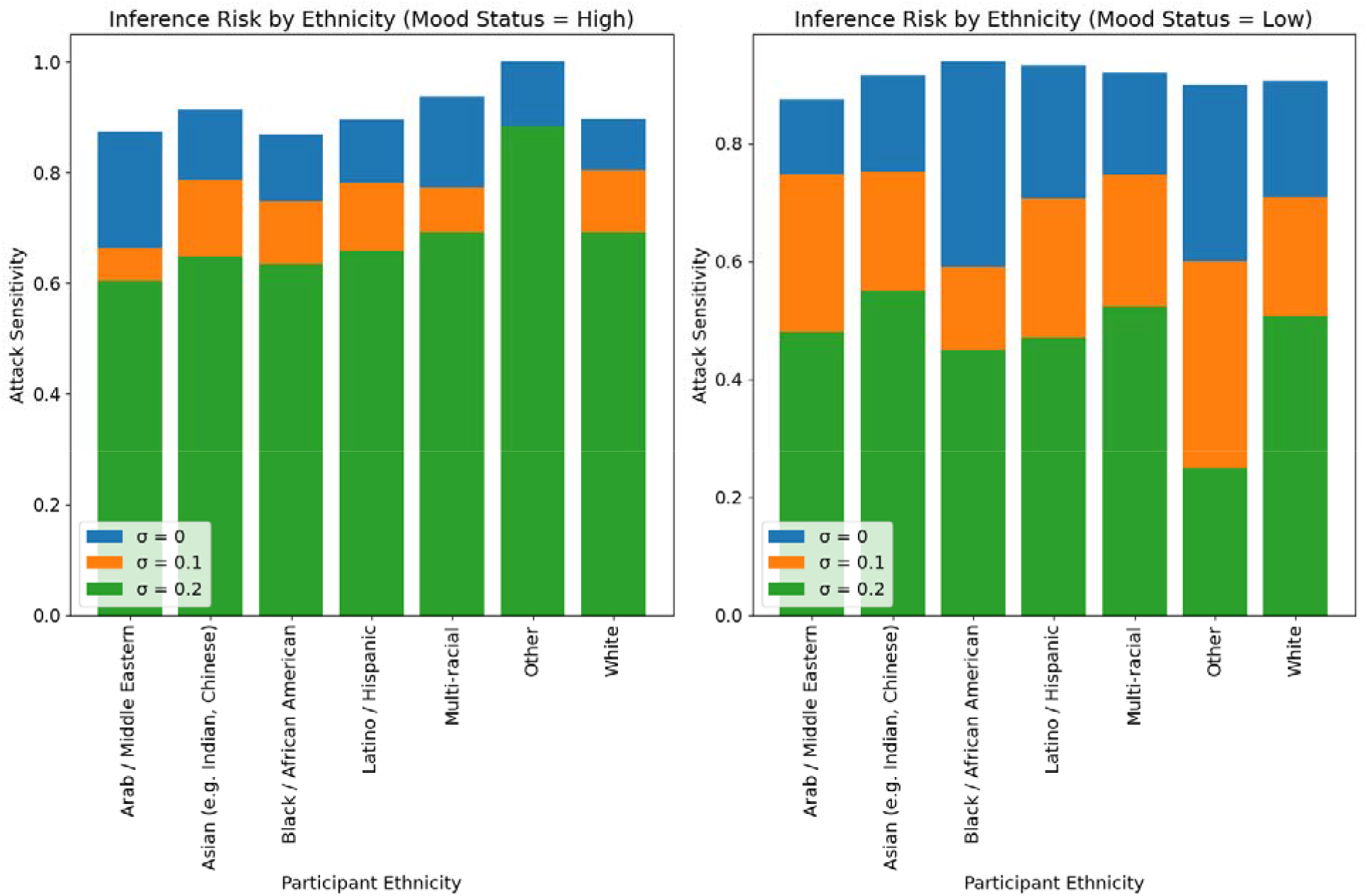
Attack Sensitivity by Ethnicity and Mood Status

## Appendix II Results from the Binary Classification Task

For the classification task in the Target System, we use the same structure and training procedure for our neural networks as the one outlined in **Appendix I** for the regression task, except we replace MSE loss with Binary Cross Entropy (BCE) loss. The construction of the External Attack does not change from the details given in **Appendix I**.

Under the binary classification task for the Target System, the attacker’s success rate in identifying private data attributes is over 80% in the worst case. However, under the highest level of DP tested in this paper, the attacker’s success rate falls to around 54.6% with only a 5 percentage point decrease in model accuracy and an apparent 40% increase in model training time.

Figure 14 plots the Target System model loss and accuracy metrics over each gradient update. Training progress under the conventional centralized training protocol is also plotted for reference. A σ value of zero denotes FL without any additional DP. We see very similar trends to those in the regression task, although we note that the model did not seem to converge within 1,000 epochs of training for the centralized regime and most values of the noise scale. This may affect our measurement of training times according to the stated definitions of convergence in the main body of the paper.

**Figure 14:**
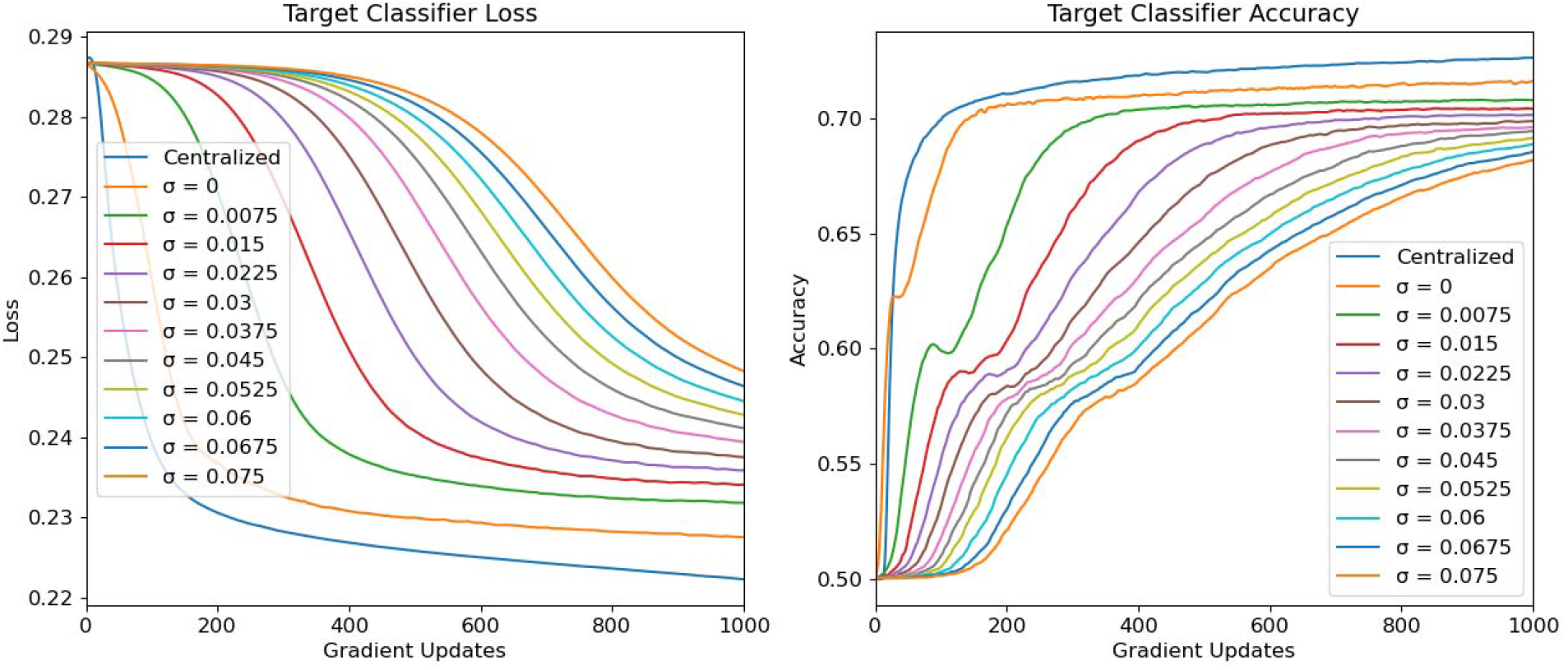
Target System Model Performance (Binary Classification Task)

Figure 15 shows the relationship between training time/final model utility and noise scale. We observe similar results as those generated in the regression task.

**Figure 15:**
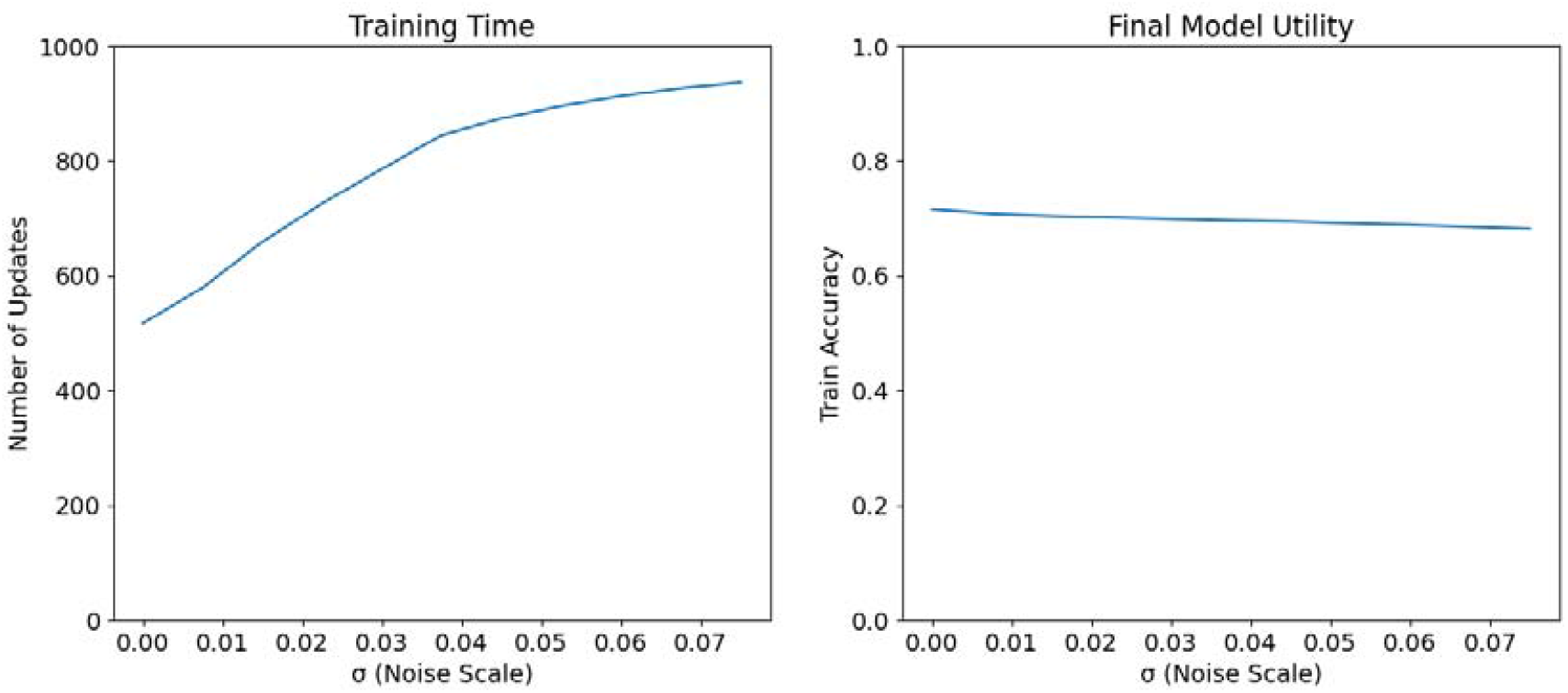
Training Time/Final Model Utility vs. Noise Scale (Binary Classification Task)

Figure 16 shows the External Attack’s success in inferring a particular participant’s mood status based on their observed gradient to the fixed Target System model parameters. We observe similar trends as those found in the regression task.

**Figure 16:**
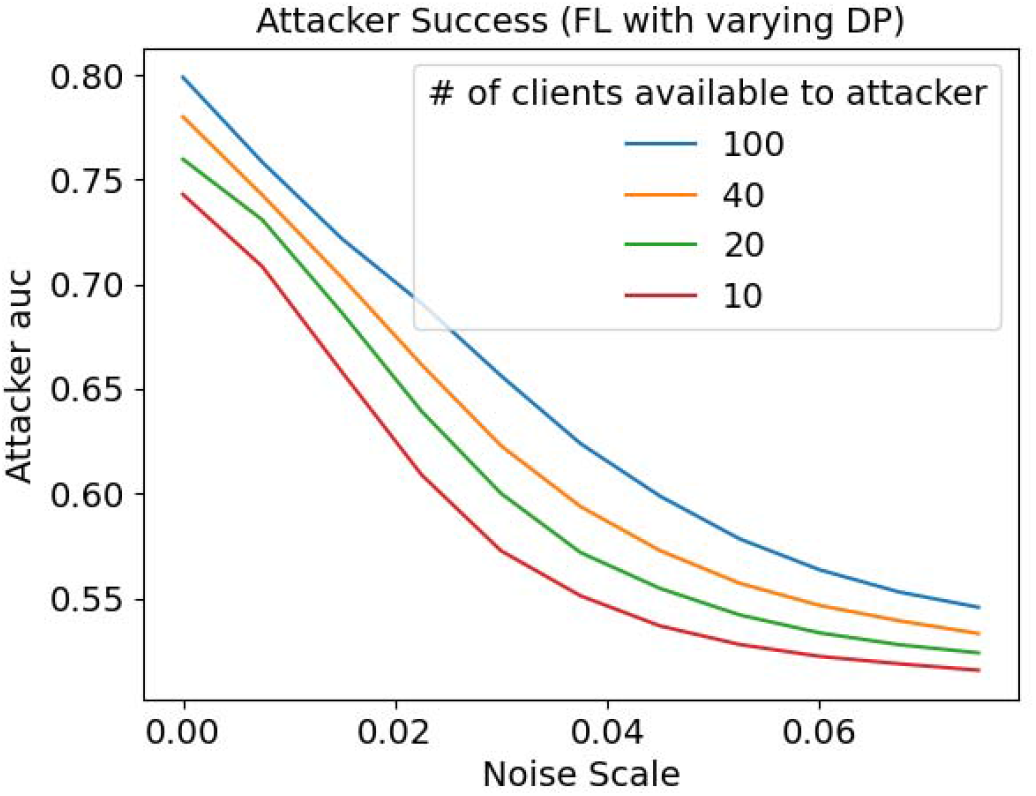
Attacker Success vs. Noise Scale (Binary Classification Task)

Figure 17 shows the property inference attack’s positive predictive value and sensitivity, given that a low mood status is viewed as a “positive test result”. We find similar results to the regression task for PPV, but we observe that trends for sensitivity across the noise scale and number of participants available to the attacker are less stable than those present in the results for the regression task. Specifically, we observe that sensitivity seems to increase at higher noise scales, although the results from attack AUC and PPV seem to suggest that the attack model may be attempting to compensate for higher noise by simply classifying more participants as “low” mood status overall.

**Figure 17:**
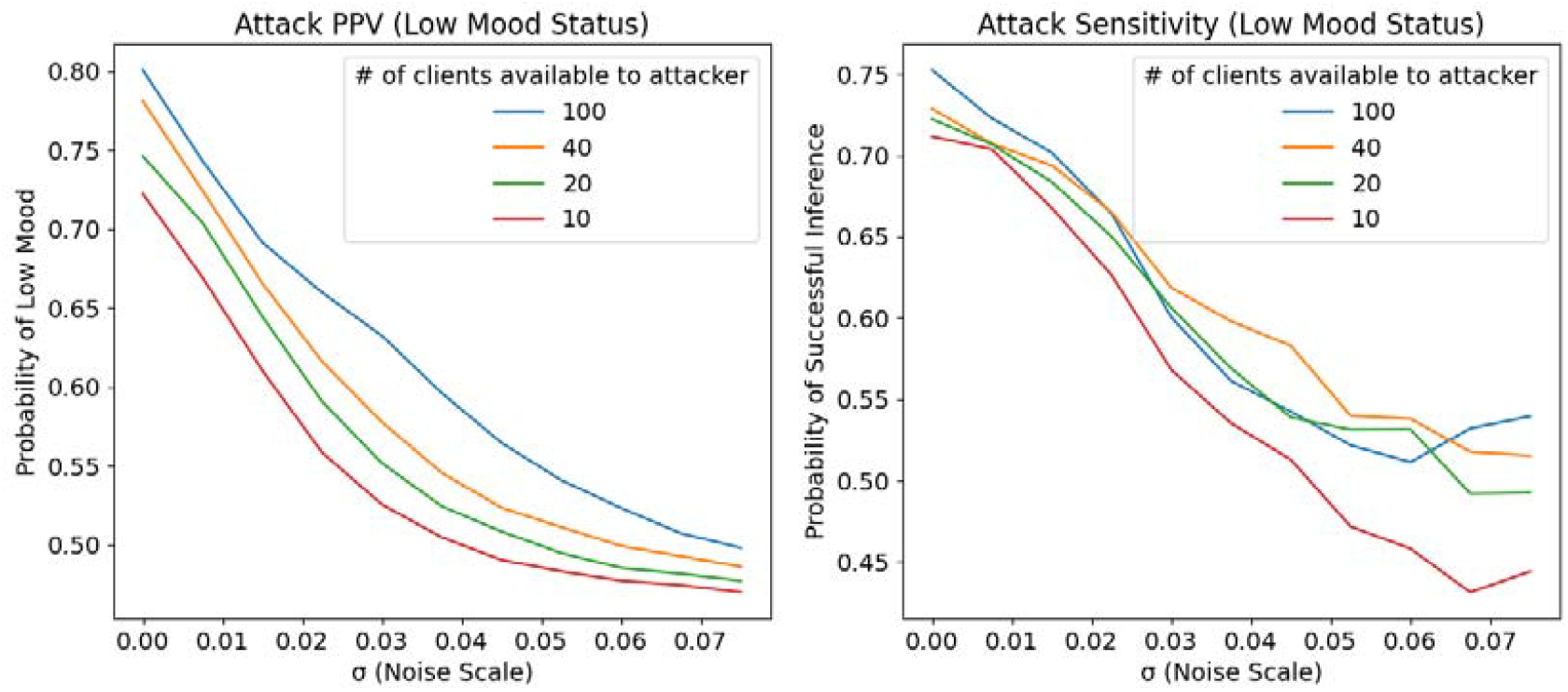
Attack PPV/Sensitivity vs. Noise Scale (Binary Classification Task)

Figure 18 shows the correlation between the attacker’s prediction of a particular participant’s mood status versus the participant’s actual mood status. We observe similar trends as those found in the results for the regression task.

**Figure 18:**
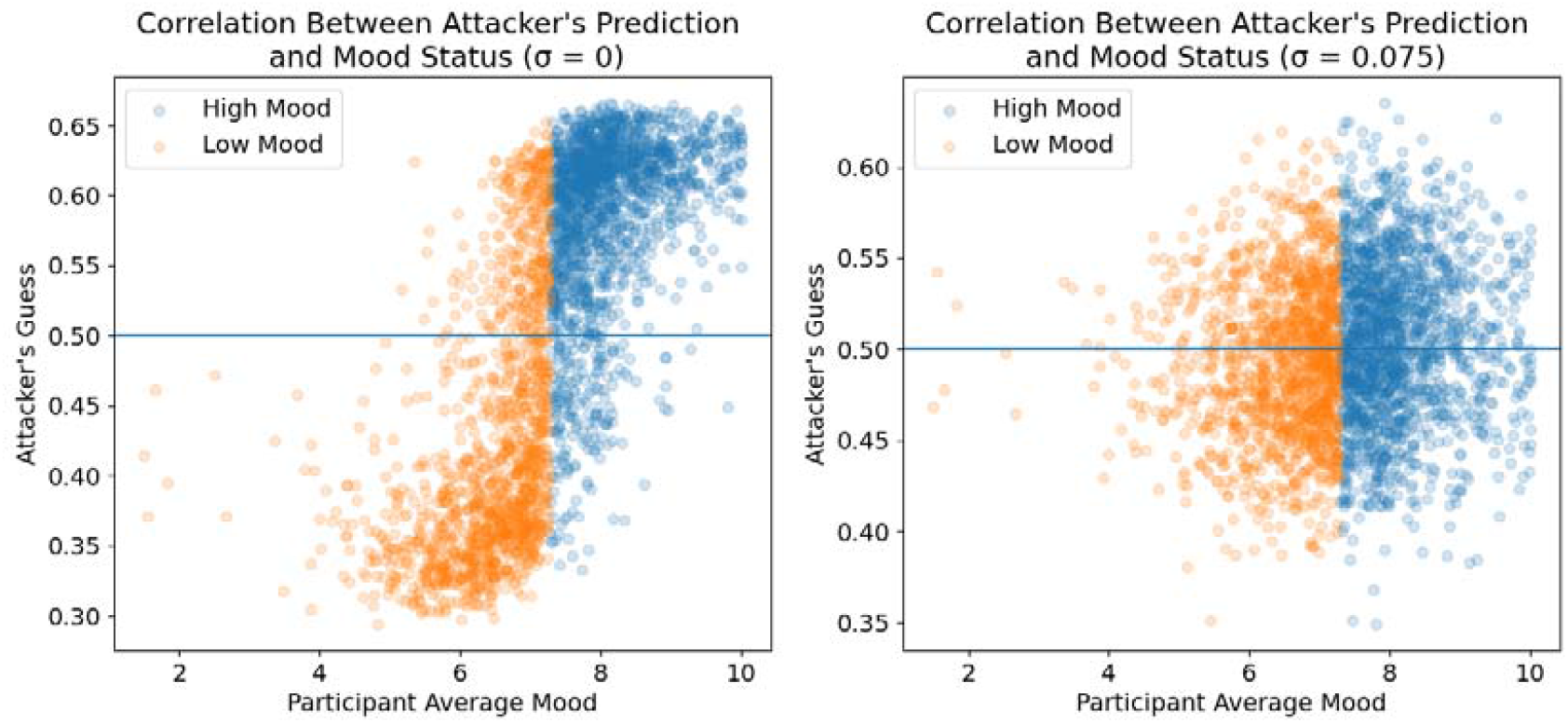
Attacker’s Predicted Mood Status vs. Participant Average Mood for Different Noise Levels (Binary Classification Task)

Figure 19 shows the External Attack’s sensitivity when the attacker has access to data for 100 IHS participants, split by mood status and age. There seem to be differences across age, although overall trends are hard to identify. We note that for some age groups in the low mood status participant block, adding noise actually slightly increases the attack sensitivity. This may be a result of the External Attack’s batch property classifier overfitting to its training data for certain demographic groups.

**Figure 19:**
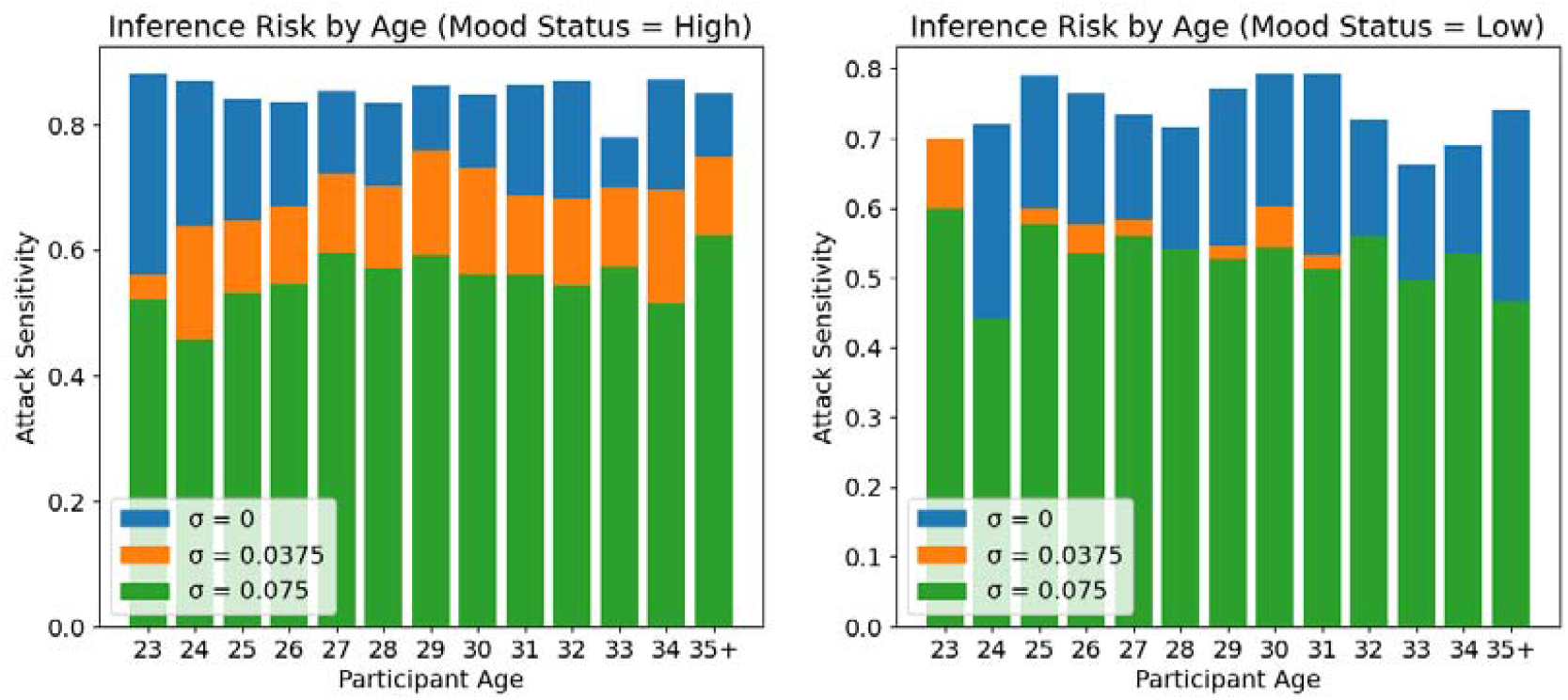
Attack Sensitivity by Age and Mood Status (Binary Classification Task)

Figure 20 shows the same data split by participant sex. There seem to be no significant differences between male and female participants.

**Figure 20:**
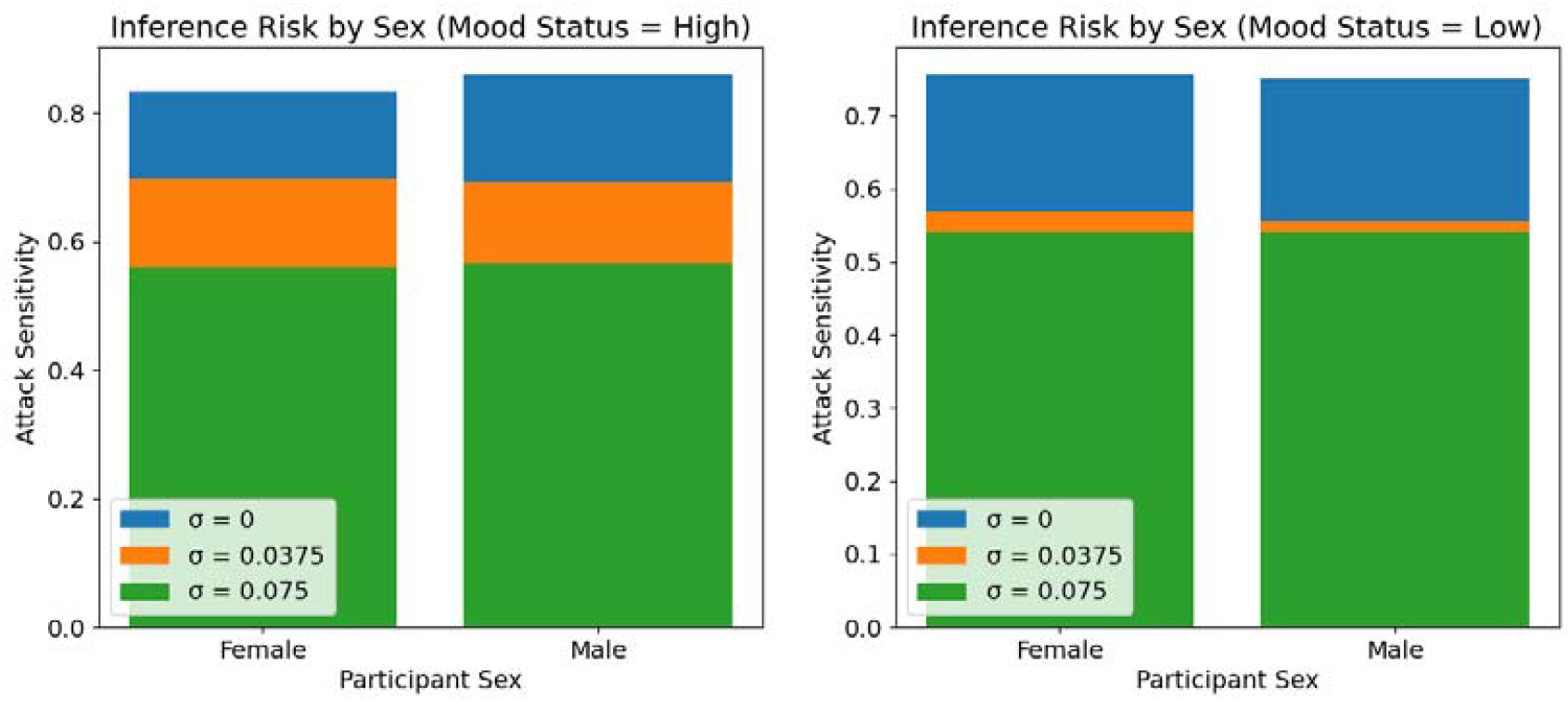
Attack Sensitivity by Sex and Mood Status (Binary Classification Task)

Figure 21 shows the same data split by participant ethnicity. For those with low mood status, adding additional noise to the Target System gradient updates seems to have heterogeneous effects on attack sensitivity, although small participant counts for some ethnic groups complicate drawing conclusions from these results.

**Figure 21:**
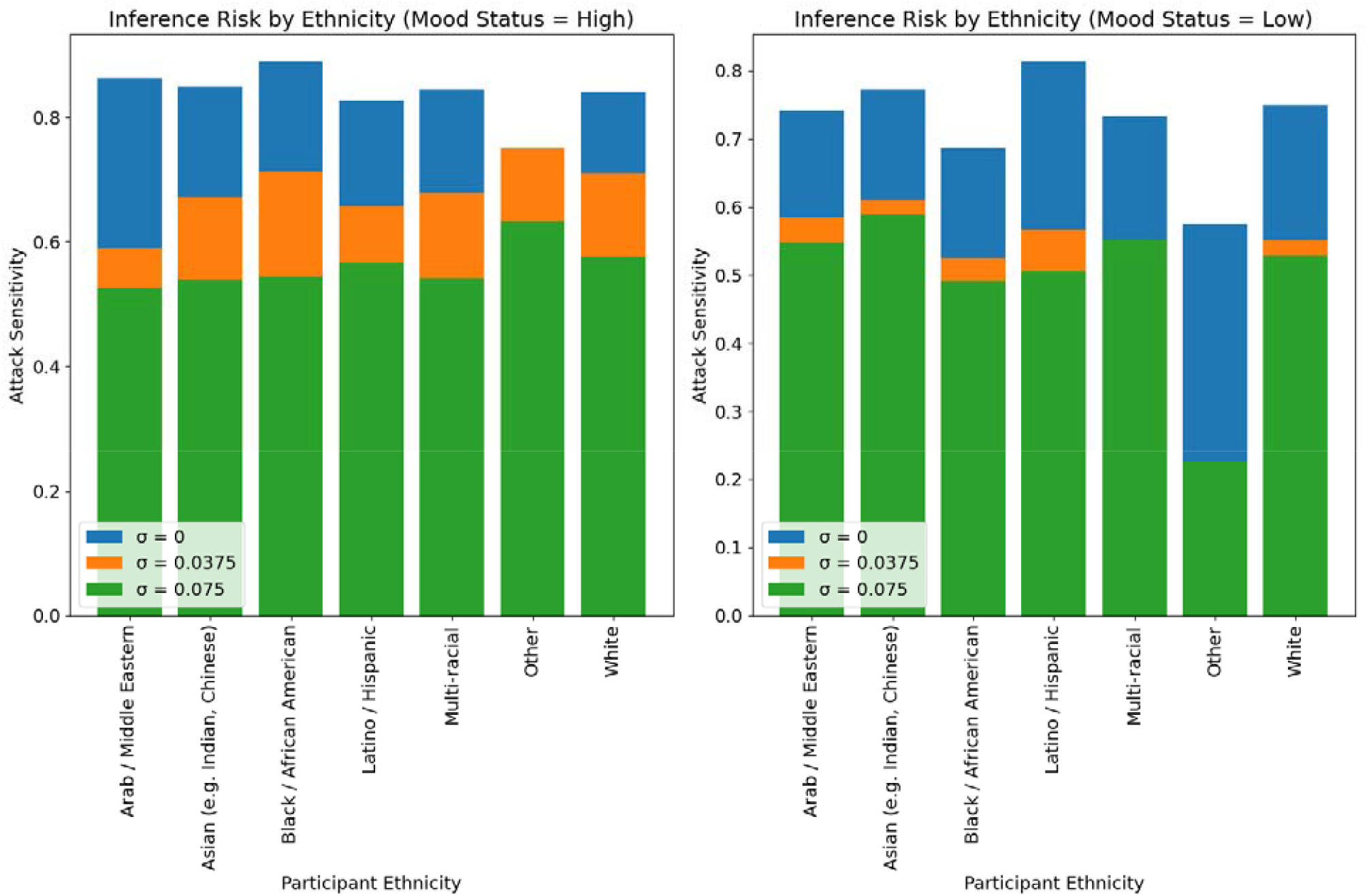
Attack Sensitivity by Ethnicity and Mood Status (Binary Classification Task)

## References

1. ltd, R. and M. Wearable Technology Market Size, Share & Trends Analysis Report By Product (Wrist-Wear, Eye-Wear & Head-Wear, Foot-Wear, Neck-Wear, Body-wear), By Application, By Region, and Segment Forecasts, 2021-2028. https://www.researchandmarkets.com/reports/5124989/wearable-technology-market-size-share-and-trends.

2. mHealth Market Size & Share, Trends Report, 2022-2030. https://www.grandviewresearch.com/industry-analysis/mhealth-market.

3. Cao, J., Lim, Y., Sengoku, S., Guo, X. & Kodama, K. Exploring the Shift in International Trends in Mobile Health Research From 2000 to 2020: Bibliometric Analysis. JMIR MHEALTH UHEALTH 17 doi:10.2196/31097.

4. Kotz, D., Gunter, C. A., Kumar, S. & Weiner, J. P. Privacy and Security in Mobile Health: A Research Agenda. Computer 49, 22–30 (2016).

5. Auxier, B. et al. Americans and Privacy: Concerned, Confused and Feeling Lack of Control Over Their Personal Information. Pew Research Center: Internet, Science & Tech https://www.pewresearch.org/internet/2019/11/15/americans-and-privacy-concerned-confused-and-feeling-lack-of-control-over-their-personal-information/ (2019).

6. Guo, X., Zhang, X. & Sun, Y. The privacy–personalization paradox in mHealth services acceptance of different age groups. Electron. Commer. Res. Appl. 16, 55–65 (2016).

7. Koffi, B., Yazdanmehr, A. & Mahapatra, R. Mobile Health Privacy Concerns - A Systematic Review. AMCIS 2018 Proc. (2018).

8. Brooks, C. 3 Key Cybersecurity Trends To Know For 2021 (and On …). Forbes https://www.forbes.com/sites/chuckbrooks/2021/04/12/3-key-cybersecurity-trends-to-know-for-2021-and-on-/.

9. Zhao, Y. et al. Federated Learning with Non-IID Data. (2018) doi:10.48550/arXiv.1806.00582.

10. Liu, J. C., Goetz, J., Sen, S. & Tewari, A. Learning From Others Without Sacrificing Privacy: Simulation Comparing Centralized and Federated Machine Learning on Mobile Health Data. JMIR MHealth UHealth 9, e23728 (2021).

11. Dwork, C. & Roth, A. The Algorithmic Foundations of Differential Privacy. Found. Trends® Theor. Comput. Sci. 9, 211–407 (2013).

12. Abadi, M. et al. Deep Learning with Differential Privacy. in Proceedings of the 2016 ACM SIGSAC Conference on Computer and Communications Security 308–318 (Association for Computing Machinery, 2016). doi:10.1145/2976749.2978318.

13. Brisimi, T. S. et al. Federated learning of predictive models from federated Electronic Health Records. Int. J. Med. Inf. 112, 59–67 (2018).

14. Lee, G. H. & Shin, S.-Y. Federated Learning on Clinical Benchmark Data: Performance Assessment. J. Med. Internet Res. 22, e20891 (2020).

15. Dayan, I. et al. Federated learning for predicting clinical outcomes in patients with COVID-19. Nat. Med. 27, 1735–1743 (2021).

16. Can, Y. S. & Ersoy, C. Privacy-preserving Federated Deep Learning for Wearable IoT-based Biomedical Monitoring. ACM Trans. Internet Technol. 21, 21:1-21:17 (2021).

17. Ficek, J., Wang, W., Chen, H., Dagne, G. & Daley, E. Differential privacy in health research: A scoping review. J. Am. Med. Inform. Assoc. 28, 2269–2276 (2021).

18. Niinimäki, T., Heikkilä, M. A., Honkela, A. & Kaski, S. Representation transfer for differentially private drug sensitivity prediction. Bioinformatics 35, i218–i224 (2019).

19. Liu, X., Zhou, P., Qiu, T. & Wu, D. O. Blockchain-Enabled Contextual Online Learning Under Local Differential Privacy for Coronary Heart Disease Diagnosis in Mobile Edge Computing. IEEE J. Biomed. Health Inform. 24, 2177–2188 (2020).

20. Choudhury, O. et al. Differential Privacy-enabled Federated Learning for Sensitive Health Data. Preprint at https://doi.org/10.48550/arXiv.1910.02578 (2020).

21. Hsu, J. et al. Differential Privacy: An Economic Method for Choosing Epsilon. in 2014 IEEE 27th Computer Security Foundations Symposium 398–410 (2014). doi:10.1109/CSF.2014.35.

22. Naseri, M., Hayes, J. & De Cristofaro, E. Local and Central Differential Privacy for Robustness and Privacy in Federated Learning. Preprint at https://doi.org/10.48550/arXiv.2009.03561 (2022).

23. Sen, S. et al. A prospective cohort study investigating factors associated with depression during medical internship. Arch. Gen. Psychiatry 67, 557–565 (2010).

24. Azur, M. J., Stuart, E. A., Frangakis, C. & Leaf, P. J. Multiple imputation by chained equations: what is it and how does it work? Int. J. Methods Psychiatr. Res. 20, 40–49 (2011).

25. Melis, L., Song, C., De Cristofaro, E. & Shmatikov, V. Exploiting Unintended Feature Leakage in Collaborative Learning. Preprint at https://doi.org/10.48550/arXiv.1805.04049 (2018).

26. Github repository for reproducibility: https://github.com/Alex-Shen-93/ihs-privacy-paper

